# Single-cell multi-omics show disruption of blood and airway T-cells in pulmonary long COVID

**DOI:** 10.1101/2025.09.18.25336101

**Authors:** Elizabeth Guinto, I-Chih Kuo, Sameeksha Chopra, Samuel B. Shin, Firoozeh V. Gerayeli, Jung-Wan Yoo, Hye-Yun Park, Melina Messing, Xuan Li, Cassandra Gilchrist, Dina Yehia, Rachel L. Eddy, Stephen Milne, Tara R. Stach, Chung Y. Cheung, Julia S.W. Yang, William Yip, Tawimas Shaipanich, Jonathon Leipsic, Kelly M. McNagny, Janice M. Leung, Don D. Sin

## Abstract

**Background:** Approximately 10% of individuals who recover from COVID-19 experience residual respiratory symptoms impacting their quality of life, but the mechanisms behind pulmonary long COVID (PLC) are largely unknown.

**Objectives:** We characterized airway and circulating immune cells in patients with and without PLC.

**Methods:** Participants were recruited and allocated into two groups: 1) PLC, defined by a St. George’s Respiratory Questionnaire (SGRQ) total score of >10 at least three months following an acute SARS-CoV-2 infection with self-reported new or worsening symptoms, and 2) controls, defined by SGRQ =<10 with or without a prior history of COVID. We performed research bronchoscopy and obtained bronchoalveolar lavage (BAL) in seven PLC patients and seven age- and sex-matched control subjects. Single-cell RNA sequencing (scRNAseq) was performed on the BAL cells. Peripheral blood mononuclear cells (PBMCs) were cryopreserved in 30 participants (17 PLC, 13 controls) for proteomic analysis. Serum was submitted for microarray detection of auto-IgG antibodies.

**Methods:** We annotated 105,836 cells using scRNAseq and found that CD4+ T-cells were credibly increased in participants with PLC. scRNAseq also revealed up-regulation of anti-viral pathways including those related to interferon signaling in T-cells as well as antigen presenting cells. In PBMCs, T-cells expressing both CD4 and CD8 were elevated in PLC participants. Autoantibodies targeting genomic DNA, collagen II, and TIF-gamma were significantly increased in PLC patients.

**Conclusions:** PLC is associated with dysregulation of T-cell mediated immunity, which may be related to autoimmunity. These cells represent potential novel therapeutic targets in patients suffering from PLC.

## Introduction

Over 700 million people worldwide have contracted SARS-CoV-2^1^. An estimated 10-30% of survivors experience persistent symptoms lasting longer than three months^2^, which has been termed ‘long COVID^3^. The impact of these respiratory symptoms on patient quality of life varies, with some experiencing symptoms severe enough to prevent them from continuing to work^4^. Respiratory symptoms, such as shortness of breath, account for approximately 24% of these cases^5^. The full impact of this disease on the healthcare system remains to be seen.

Currently, the underlying cellular mechanisms driving the pulmonary symptoms in long COVID are largely unknown. Previous studies have focused largely on hospitalized individuals and used peripheral blood to identify genomic or proteomic signatures of the disease ^6–8^. As SARS-CoV-2 is a respiratory virus that first infects the nasal epithelium, the airway mucosa is among the first lines of defense against the initial viral infection^9^. Studies have shown that long COVID patients have persistent radiological abnormalities as a surrogate for the impacts of COVID-19 infections related to the long COVID on the airways^10,11^. Despite this, very few studies have directly interrogated the airways for insights on disease mechanisms or biomarker^12^. Long COVID is a heterogeneous disease, which encompasses many symptoms, severities, and timelines^2^. It is more common in young women and is not limited to those who had a severe acute infection of SARS-CoV-2^2^.

Single-cell RNA sequencing (scRNAseq) is a powerful tool to characterize cell populations and determine changes in gene expression across and within populations^13^. It has been used to identify specific cell subtypes and characterize a heterogeneous population of cells within a tissue^13^. Unlike antibody-based proteomic analyses, scRNAseq is unsupervised and exploratory and thus does not present the same biases^13^. However, the relationship between mRNA and protein is not always linear^14^. Proteomic technologies validate transcriptomic-based assays as proteins ultimately dictate cellular function. One method to assess proteomic signatures is mass cytometry or cytometry by time-of-flight (CyTOF), which can detect over thirty antigens per sample using metal isotopes rather than fluorescent tags. This feature maximizes available markers by eliminating the need for compensation of fluorescent spectral overlap^15^. In effect, this approach acts as a single-cell proteomic tool for >30 markers. Here, we built on previous work and scRNAseq and CyTOF technologies to describe the cellular landscape of the airway and peripheral immune cells of patients with pulmonary long COVID (PLC) and provide mechanistic insights on its pathophysiology^16,17^.

## Methods

### Participant Recruitment

We used data from patients with long COVID^18^, who had enrolled in the Biomarker Discov<u>e</u>ry for the Po<u>st</u>-Covid <u>P</u>ulmonary <u>S</u>yndrom<u>e</u> (RESPONSE) cohort, which has been previously described in detail^12^. Control subjects were also recruited from the Canadian Users of Cannabis Smoke (CANUCK) Study cohort; only subjects without a history of cannabis or tobacco cigarette smoking were included. The inclusion and exclusion criteria can be found in the <u>Supplementary Materials</u>. Subjects with pre-existing severe pulmonary disease were not included, and participants with previous SARS-CoV-2 infections had to have been three or more months post-infection at the time of enrolment. Participants were recruited who were unexposed to SARS-CoV-2, had an exposure to SARS-CoV-2 but fully recovered without residual respiratory symptoms or had an exposure to SARS-CoV-2 with new or worsening pulmonary symptoms >12 weeks post-infection assessed by the Saint George’s Respiratory Questionnaire (SGRQ), which is a pulmonary-specific health status instrument^19^. An SGRQ total score >10 denoted pulmonary long COVID (PLC) as previously defined and persons with a score below this threshold were considered non-symptomatic and placed in the control group^18^. Blood was collected from these participants, as described in the <u>Supplementary Materials</u>, from which peripheral blood mononuclear cells (PBMCs) and serum were isolated for subsequent analyses.

### Bronchoalveolar Lavage acquisition and processing

Consenting participants underwent bronchoscopy as previously described^20^. After collection, the bronchoalveolar lavage (BAL) sample was processed using methods reported by Gerayeli et. al.^21^. Once processed, the cells were transferred on ice to the Biomedical Research Centre at the University of British Columbia for library preparation and sequencing using the 10X Genomics platform. The transcriptome was mapped to the human genome, version hg38. Cells were also submitted for bulk RNA-sequencing, as described in the <u>Supplementary Materials.</u>

### Cytometry by time-of-flight

The CyTOF samples were prepared according to the method described by Messing et. al^22^ and subsequently evaluated using the Helios CyTOF2^®^ mass cytometer, with a maximum event captured threshold set to 500,000 events per sample at a flow rate of 30μL/min. Of the 38 surface proteins incorporated into the panel, 22 of them were cell-type markers (such as TCR-αβ and CD3ε for T-cells) and the remaining 16 were cell-state markers. Panel validation was performed as previously described^23^. The full antibody list is available in Supplementary Table 1 (PBMCs) and 2 (BAL).

### Serum Antibody Assays

Serum was submitted for testing of 120 IgG autoantibodies at the University of Texas Southwestern Medical Center as previously described by Rojas et. al.^24^. All participants with PLC had a positive result on their Roche® Anti-N antibody test, indicating that an antibody response against SARS-CoV-2 was maintained many months post-infection; the assay is described in the <u>Supplementary Materials</u>.

### Data Analysis

#### Single-cell sequencing

The scanpy package for Python was used for quality control, batch correction, dimensionality reduction, and Leiden clustering^25^. The clusters were annotated both manually (Supplementary Table 3) and with the assistance of the CellTypist^26^ algorithm, using the majority votes feature. Gene expression and pathway analysis was performed using the GseaPy^27^ package for gene set enrichment analysis (GSEA) against the Reactome^28^ 2022 pathway database. The decoupler package for python was used for pseudobulk differential gene expression analysis^29^. Briefly, pseudobulk analysis pools all cells of the same type (ex. CD4+ T-cells) from the same participant and adds the gene expression between all cells to create a gene expression profile. This way, each participant is counted as one sample as opposed to each cell being its own sample. Cell proportions from scRNAseq were analyzed using the sc-CODA^30^ Bayesian algorithm. Bayesian models do not use p-values but rather generate data using a probability-based model. The SC-CODA model uses one cell type as a reference and revaluates the other cell types based on the assumption that the reference is unchanged between the groups. In this framework, we used each cell type as a reference and considered cell types that were increased ≥ 50% of the time to be statistically credible. Cell interactions were evaluated using the CellPhoneDB package statistical analysis method^31^. Further details are provided in the <u>Supplementary Materials</u>.

#### CyTOF

Quality control was performed using the premessa package in R^32^ and FlowJo gating software. Batch correction was done using CyCombine, a combat-based algorithm^33^. BAL samples were all batched together in one run, thus did not require batch correction. Further analysis, including clustering and dimensionality reduction, was performed by using the FlowKit and scanpy packages for Python as previously described^23,25,34^. Annotation was performed manually based on established markers. Cell proportion differences between control and PLC participants were assessed using a Wilcoxon’s rank sum test. Further detail is provided in the <u>Supplementary Materials.</u>

## Results

### Bronchoscopy cohort demographics

We obtained BAL samples for scRNAseq from 14 participants: seven PLC patients, and seven controls matched for age, sex, and smoking history (Figure 1). All participants tested negative for COVID-19 before undergoing bronchoscopy and all participants had normal lung function tests (PFTs) and no clinically significant computed tomography (CT) abnormalities. All PLC participants had mild (non-hospitalized) bouts of acute COVID-19. No significant difference in concentrated BAL supernatant SARS-CoV-2 N protein concentration was detected (p=0.65). Of the seven controls, five subjects did not have a previous SARS-CoV-2 infection. One PLC participant had two BAL samples sequenced, and both were used for the analysis. Demographics, differential BAL cell count, and qualitative CT scan data are shown in Table 1. There were no significant differences in the qualitative CT or differential cell composition in the BAL sample between the PLC and control participants. In both groups, alveolar macrophages were the most abundant cell type, accounting for ∼80% of the cells, which was followed by lymphocytes (∼10%); there was a scarcity of granulocytes in the BAL (<5%). PLC patients were 567 ± 269 days between their bout of acute COVID-19 and their bronchoscopy while controls were 521 ± 314 days post-COVID-19 (p=0.9). PLC participants were persistently symptomatic from time of infection to time of bronchoscopy.

**Figure 1.**
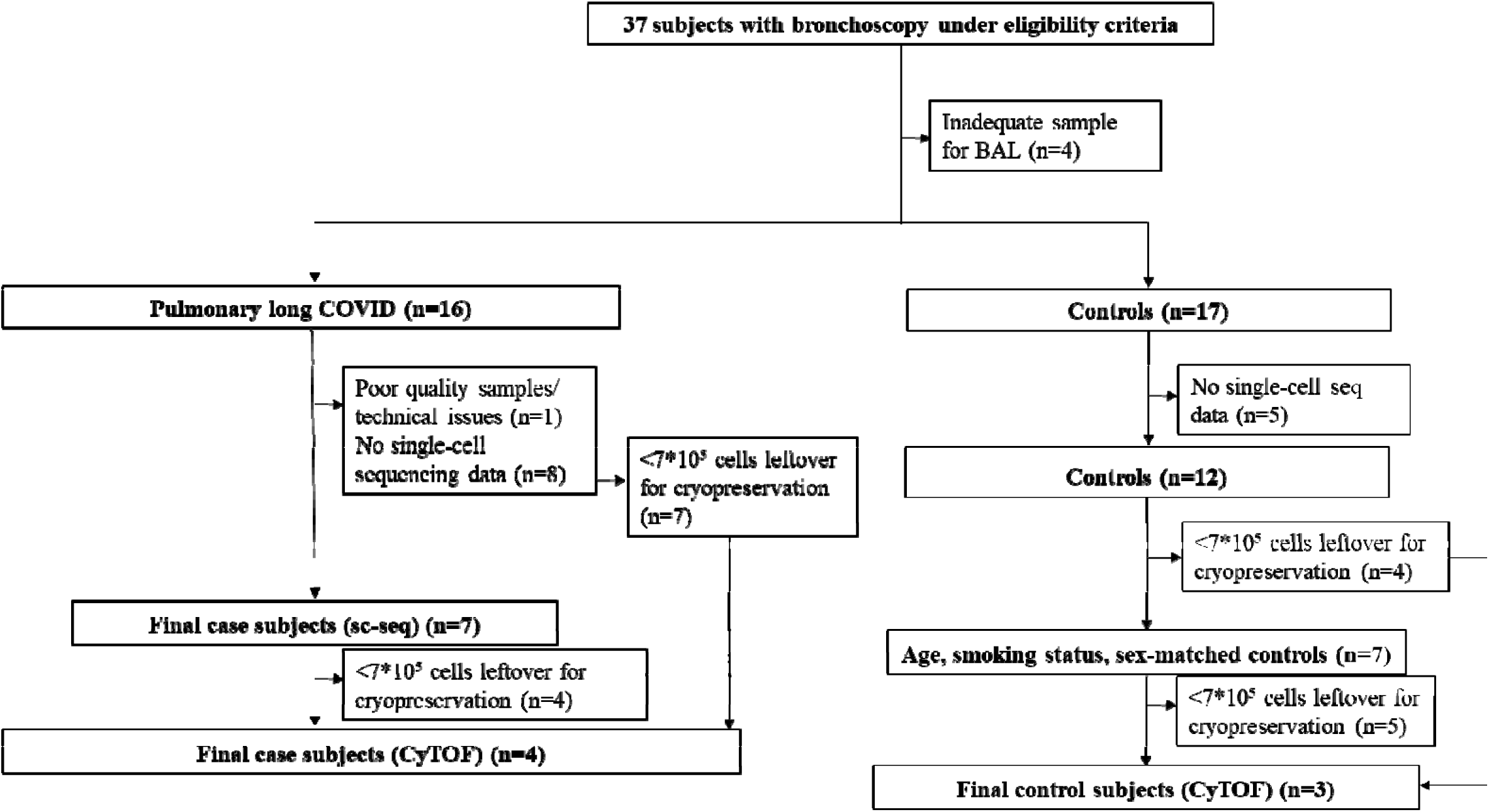
A flowchart depicting the research bronchoscopy cohort and the selection of the participants. Participants with pre-existing severe lung conditions, such as chronic obstructive pulmonary disease and severe asthma were excluded, alongside heavy smokers (both cigarette and cannabis) and e-cigarette users. If the bronchoalveolar lavage has considerable blood contamination (>30%) or the sample sequencing failed, this was considered poor sample quality/technical issues.

**Table 1.** Participant demographics for single cell sequencing of bronchoalveolar lavage (BAL) cells. All values are expressed as mean [standard deviation] unless otherwise specified.

|  | Pulmonary long COVID (n=7) | Control (n=7) | P-value |
| --- | --- | --- | --- |
| Age, years | 42.3 ± 10.5 | 44.0 ± 17.6 | 0.9 |
| Male, n (%) | 4 (57%) | 4 (57%) | 1 |
| BMI, kg/m<sup>2</sup> | 26.6 ± 3.4 | 25.6 ± 2.5 | 0.5 |
| Tobacco smokers, n (%) |  |  | 0.6 |
| Former | 1 (14.3%) | 2 (28.6%) |  |
| Never | 6 (85.7%) | 5 (71.4%) |  |
| Cannabis (%) |  |  | 1 |
| Never | 7 (100%) | 7 (100%) |  |
| Current | 0 (0%) | 0 (0%) |  |
| Electronic Cigarettes, n (%) |  |  | 1 |
| Never | 7 (100%) | 7 (100%) |  |
| Current | 0 (0%) | 0 (0%) |  |
| SGRQ, total | 50.6 ± 20.9 | 3.0 ± 4.4 | 0.002 |
| Symptoms | 53.4 ± 20.0 | 6.7 ± 7.0 | 0.002 |
| Activity | 63.0 ± 30.3 | 6.0 ± 10.4 | 0.003 |
| Impact | 42.3 ± 19.0 | 0.0 ± 0.0 | 0.001 |
| Pulmonary Function Test |  |  |  |
| FVC, L | 4.9 ± 0.65 | 4.7 ± 0.48 | 0.4 |
| FVC, % of predicted | 117 ± 8 | 121 ± 7 | 0.3 |
| FEV<sub>1</sub>, L | 4.0 ± 0.62 | 3.8 ± 0.43 | 0.3 |
| FEV<sub>1</sub>, % of predicted | 117 ± 6 | 118 ± 11 | 0.8 |
| FEV<sub>1</sub>/FVC | 82.2 ± 5.5 | 80.0 ± 4.2 | 0.5 |
| DLco, % of predicted | 107 ± 15 | 110 ± 11 | 0.5 |
| Blood cell counts |  |  |  |
| White blood cells (x 10<sup>9</sup>/L) | 5.4 ± 1.9 | 4.9 ± 1.0 | 1 |
| Neutrophils (%) | 53.6 ± 11.0 | 55.6 ± 5.4 | 1 |
| Lymphocytes (%) | 35.7 ± 10.2 | 33.3 ± 5.0 | 0.8 |
| Monocytes (%) | 5.3 ± 1.7 | 5.2 ± 1.2 | 0.9 |
| Eosinophils (%) | 2.1 ± 1.1 | 2.5 ± 1.4 | 0.4 |
| Bronchoalveolar Lavage |  |  |  |
| Total Nucleated cells | 158.7 ± 73.4 | 196.1 ± 167.6 | 0.9 |
| Alveolar Macrophage (%) | 84.8 ± 11.3 | 83.5 ± 9.9 | 0.7 |
| Lymphocytes (%) | 11.4 ± 9.1 | 11.0 ± 8.4 | 0.9 |
| Neutrophils (%) | 3.2 ± 3.3 | 4.4 ± 4.6 | 0.6 |
| Eosinophils (%) | 0.0 ± 0.0 | 0.29 ± 0.49 | 0.2 |
| Bronchial lining cells (%) | 0.67 ± 1.2 | 0.71 ± 1.9 | 0.1 |
| Acute COVID-19 Severity, n (%) |  |  | 0.4 |
| Hospitalized | 0 | 1 (14.3%) |  |
| Mild | 7 (100%) | 1 (14.3%) |  |
| Asymptomatic | 0 | 0 (0%) |  |
| Unexposed | NA | 5 (71.4%) |  |
| Computed Tomography, n (%) |  |  |  |
| Bronchiectasis (mild) | 2 (29%) | 0 (0%) | 0.20 |
| Consolidations | 0 (0%) | 0 (0%) | 1 |
| Ground Glass Opacities | 0 (0%) | 0 (0%) | 1 |
| Honeycombing | 0 (0%) | 0 (0%) | 1 |
| Linear scars or atelectasis | 2 (29%) | 0 (0%) | 0.20 |
| Mosaic attenuation | 0 (0%) | 0 (0%) | 1 |
| Reticulations (mild) | 1 (14%) | 0 (0%) | 0.4 |
| Emphysema | 0 (0%) | 0 (0%) | 1 |
| Vaccination status, n (%) |  | (n=3)* |  |
| Vaccinated prior to infection | 5 (71%) | 1 (50%) (n=2) | 1 |
| Received first dose prior to enrolment | 7 (100%) | 3 (100%) | 1 |
| Received second dose prior to enrolment | 7 (100%) | 3 (100%) | 1 |
| Unvaccinated at time of enrolment | 0 (0%) | 0 (0%) | 1 |

Abbreviations: BMI (body mass index), SGRQ (Saint George's Respiratory Questionnaire) FVC (forced vital capacity), FEV1 (forced expiratory volume in one second), DLco (diffusion capacity for carbon monoxide)
\*Vaccination date only available in three of the control participants

None of the samples contained residual SARS-CoV-2 mRNA based on transcriptomic mapping to the viral transcriptome. Seven BAL samples (six of which were also sequenced) contained sufficiently high number of cells to perform CyTOF, three of which were controls. The results are summarized in Supplemental Table 4.

### Single-Cell Sequencing of the BAL cells

We sequenced 105,836 cells from BAL samples, which met quality control requirements. We identified 19 different cell groups in our samples (Figure 2A). The vast majority of cells were M2 alveolar macrophages. We also found clusters of M1 alveolar macrophages, CCL3+ alveolar macrophages, metallothionein-expressing alveolar macrophages, monocyte-derived macrophages, and proliferating alveolar macrophages. In terms of lymphocytes, B-cells, CD4 T-cells, CD8 T-cells, regulatory T-cells, proliferating T-cells, and natural killer (NK) cells were all detected as were four types of dendritic cells (DCs): type 1, type 2, migratory and plasmacytoid.

**Figure 2.**
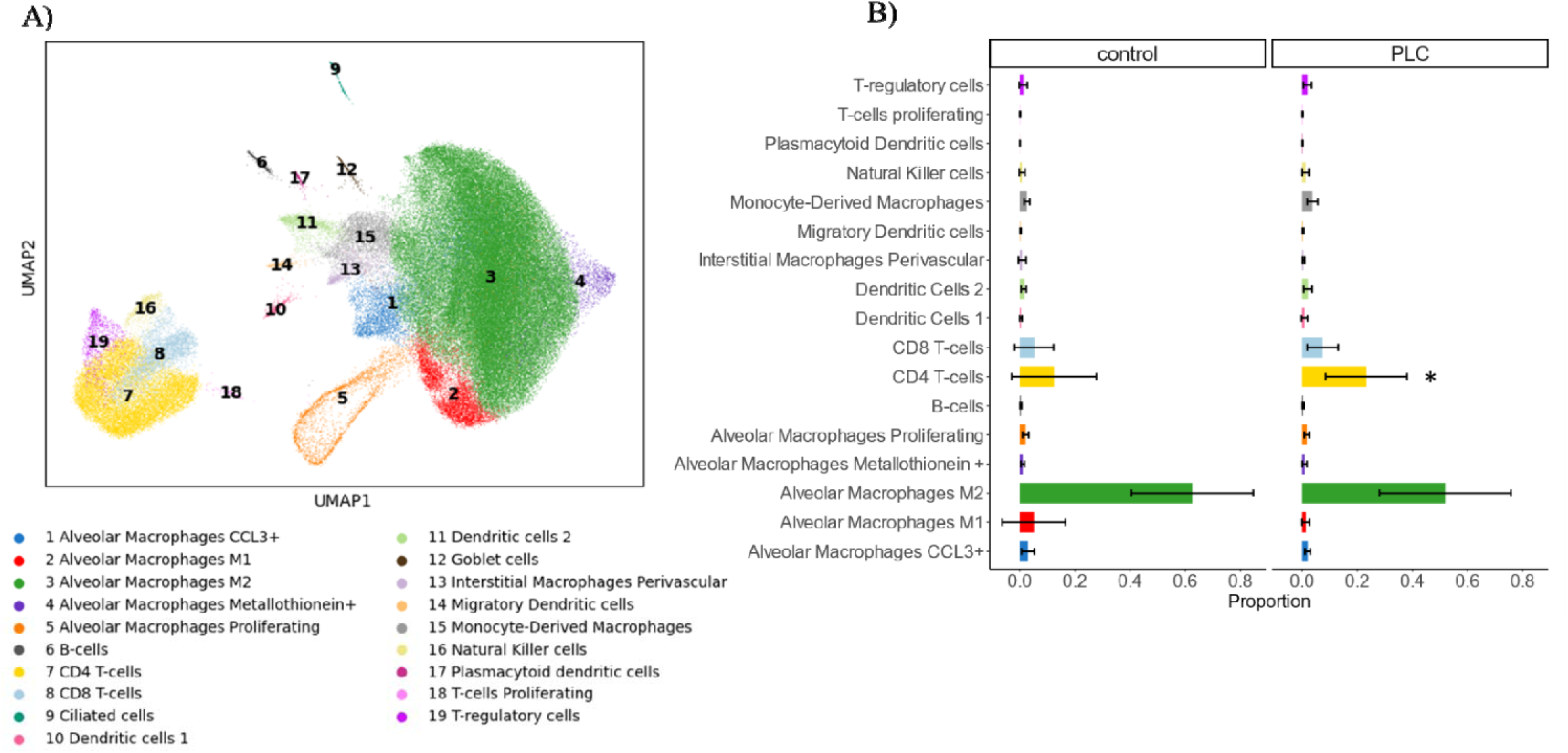
**A)** Uniform manifold approximation and projection for dimension reduction (UMAP) representation of cells in BAL based on single-cell RNA sequencing data. Cells are clustered by cell type, denoted by the colours. **B)** Composition plot showing the average cell proportions in control (left) and PLC (right) participants. Alveolar macrophages of the M2 phenotype were the most represented among both groups. CD4+ T-cells were considered statistically increased in PLC.

Most cell type proportions were comparable between the PLC and control groups. However, CD4+ T-cells were credibly increased based on the sc-CODA algorithm. Overall, there was a general shift towards a reduction in M2 alveolar macrophages and an increase in T-cells in the BAL of PLC patients (Figure 2B). Absolute counts did not differ between PLC and control participants (Supplemental Figure 1). When the PLC cohort was stratified into tertiles based on participants’ SGRQ scores, participants in the third (highest score) tertile exhibited the highest proportion of CD4+ T-cells, followed by the second and first tertiles, although this difference was not statistically significant (Supplemental Figure 2). Notably, there was a significant positive correlation between SGRQ scores and CD4+ T-cell proportions in PLC patients (R=0.83, p=0.021), as shown in Supplemental Figure 3. Additionally, CyTOF of the BAL revealed a non-significant increase of CD4+ T-cells in PLC patients (Supplemental Figure 4).

Differential gene expression and pathway analysis were performed on T-cells (including CD4+ T-cells, CD8+ T-cells, T-regulatory cells and proliferating T-cells), alveolar macrophages (including M2, M1, CCL3+, Metallothionein+ and proliferating alveolar macrophages) and DCs (including DC1s, DC2s, migratory DCs and plasmacytoid DCs) (Figure 3A). In PLC participants, all three of these cell types demonstrated increased expression of *MTRNR2L12*-a paralog of protein-coding gene. Both DCs and alveolar macrophages exhibited elevated levels of the HLA-DRB5 marker. A variety of interferon (IFN)-related genes including interferon-stimulated genes (ISGs), interferon-induced protein with tetratricopeptide repeats (IFITs) and interferon-induced transmembrane proteins (IFITMs) were upregulated in PLC samples compared to controls. Specifically, alveolar macrophages showed an increase in gene expression of *ISG15*, *IFIT2*, *IFIT3*, and *IFITM2*. Plasmacytoid dendritic cells showed higher gene levels of *ISG15*, *IFITM2*, and *IFITM3*. A pseudobulk analysis revealed that CD4+ T-cells in particular displayed increased CCL4 expression (Supplemental Figure 5).

**Figure 3.**
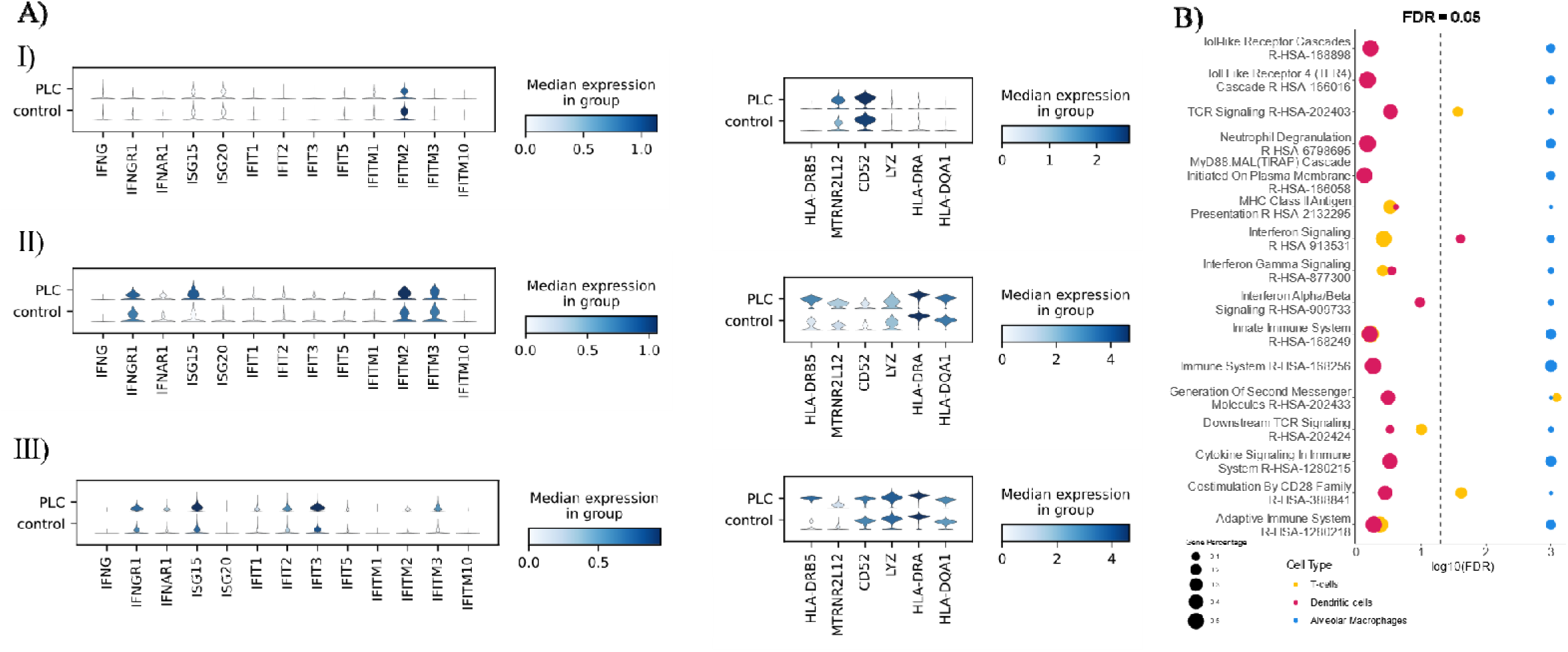
**A)** Violin plots denoting gene expression of <u>I)</u> T-cells, <u>II)</u> Dendritic cells, <u>III)</u> Alveolar Macrophages in PLC (top) and controls (bottom). Darker colours indicate stronger gene expression and large violins indicate a higher proportion of cells expressing this gene. **B)** Dot plot representing the various pathways (y-axis) enriched in PLC participant. Dots are labelled by colour to indicate which cell type they belong to, and their size indicates the proportion of genes that overlap with that pathway. The x-axis is the -log10 of the FDR for that given cell type and pathway. Abbreviations: IFN (Interferon), IFNGR (Interferon-gamma receptor), ISG (interferon-induced gene), IFIT (interferon Induced proteins with Tetratricopeptide repeats), IFITM (interferon-induced transmembrane protein), HLA (human leukocyte antigen), TCR (T-cell receptor), MHC (Major histocompatibility complex)

The results of the GSEA are presented in Figure 3B. All three cell types exhibited enrichment in immune-related pathways in PLC. Alveolar macrophages had the most significantly (FDR < 0.05) upregulated inflammatory pathways. Meanwhile, T-cells pathways were enriched for TCR-signaling, generation of second messenger molecules, and stimulation by CD28 family of cells. DCs only had one significantly upregulated pathway, which was related to interferon signaling.

We also performed cell:cell interaction analysis using the CellPhoneDB algorithm, which determines the number of interactions based on ligand and receptor expression (Figure 4)^31^. CD8+T-cells in PLC patients interacted significantly with other T-cells (including CD4+ T-cells and proliferating T-cells), natural killer cells, and alveolar macrophage subtypes. Most of these interactions were between chemokine ligands and their receptors, including *CCL2*/*CCR2*, *CCL17*/*CCR4*, *CCL5*/*CCR4*, *CCL4*/*CCR5*, *CCL5*/*CCR5* as well as tumour necrosis factor (TNF) ligand and receptor (*TNF*/*TNFRSF1B*) (Supplemental Figure 6). Overall, the analysis demonstrated a transcriptional shift in interactions from maintenance (M2 macrophages) to cellular immunity (CD8+ T-cells). Cell interactions in control participants primarily involved alveolar macrophages (M2).

**Figure 4.**
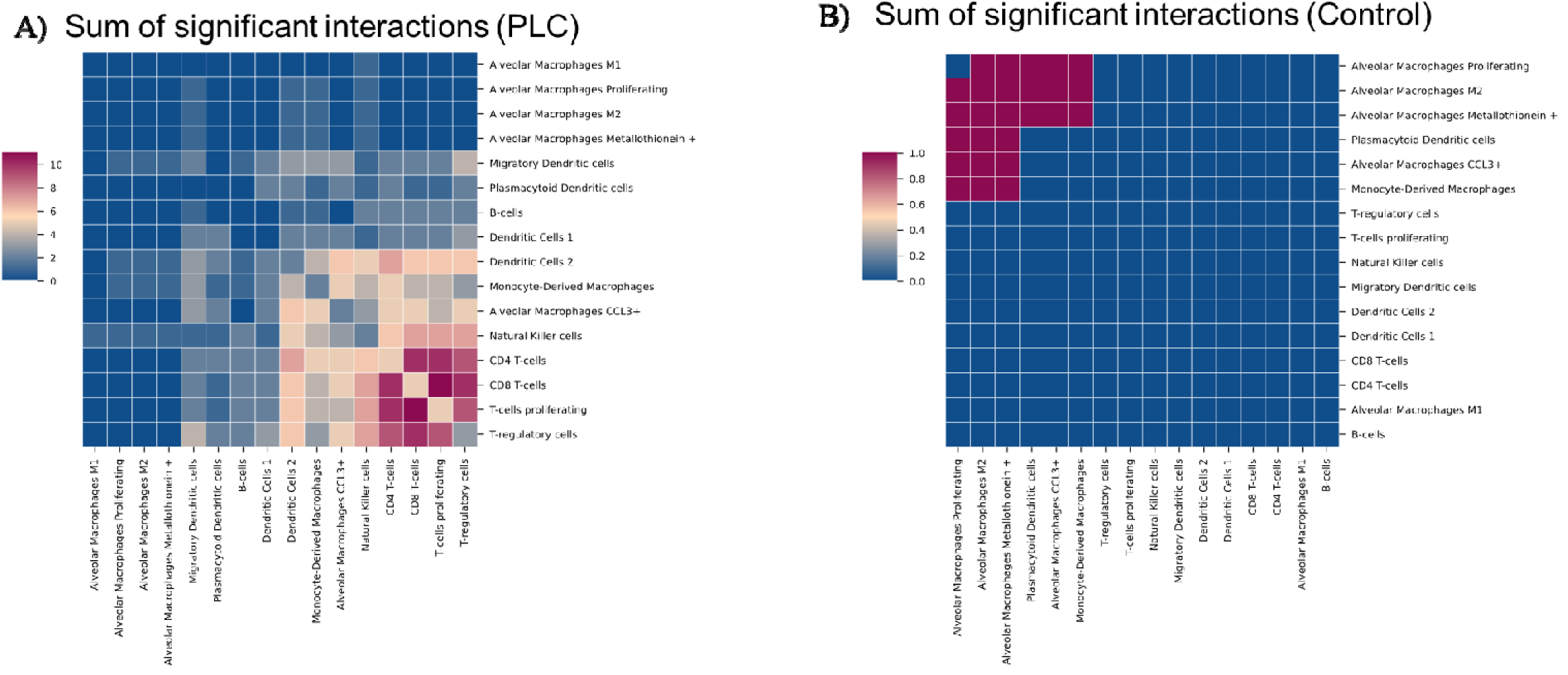
Heatmap depicting cell interactions in **A)** PLC participants and **B)** control participants. A brighter colour indicates more significant interactions between those two cell types.

### Peripheral blood mononuclear cell acquisition and demographics

Blood was obtained from study participants using standard venipuncture techniques; from which serum samples were prepared in 22 patients with PLC and 18 control participants. In a subset of these participants (n=30; 17 with PLC and 13 controls), PBMCs were isolated, cryopreserved, and prepared for CyTOF experiments. Demographic data of the study participants are summarized in Table 2. On average, PLC patients were 470 ± 241 days between their initial infection and their blood draw and controls were 363 ± 284 days post-COVID (p=0.8). All participants had normal pulmonary function test results. As with the BAL participants, the PLC group was persistently symptomatic from time of infection to time of blood draw. No participant had significantly abnormal lung function or CT scans and most participants were mild (home-recovered). Serum SARS-CoV-2 N protein was not significantly different between controls and PLC participants (p=0.65).

**Table 2.**
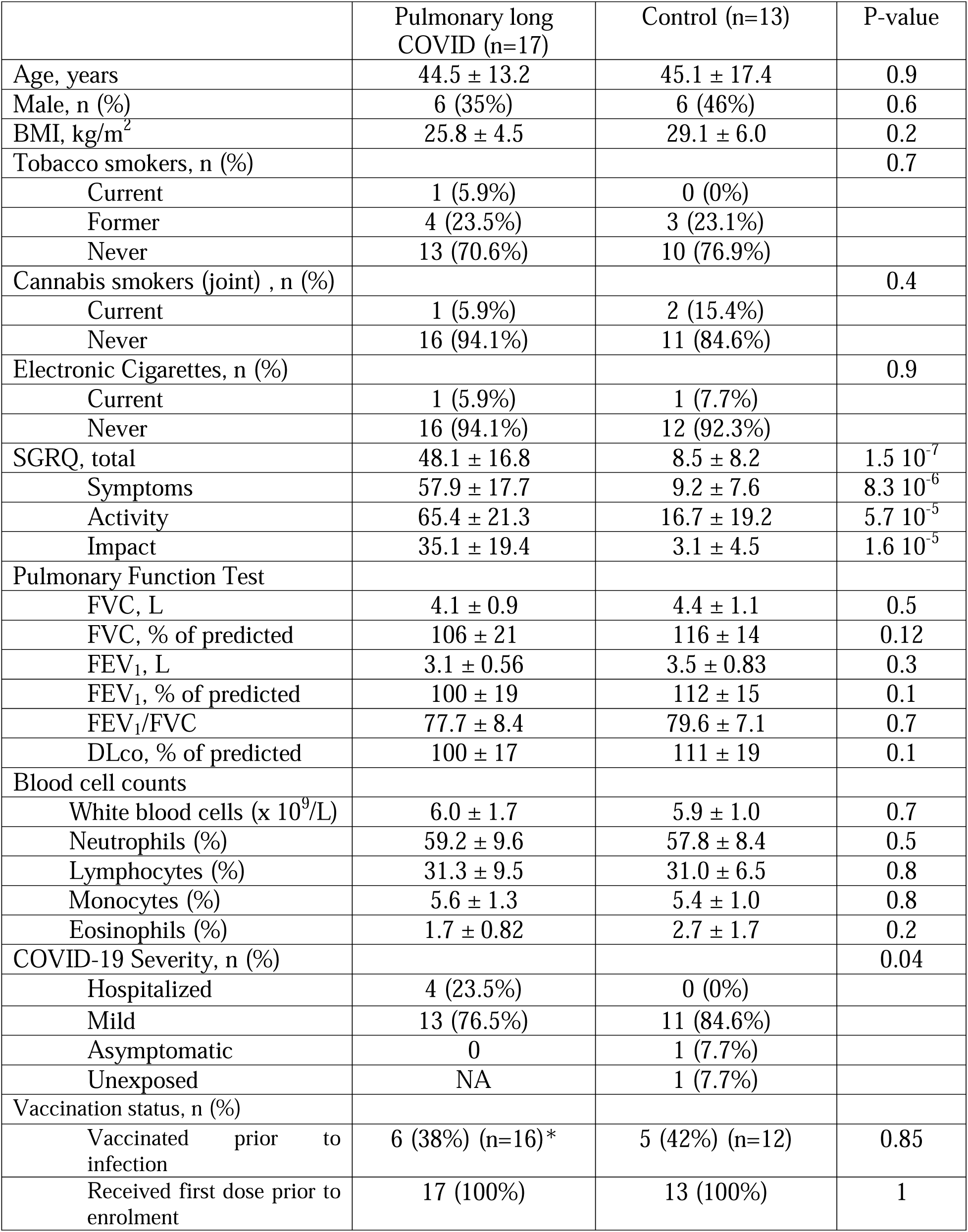
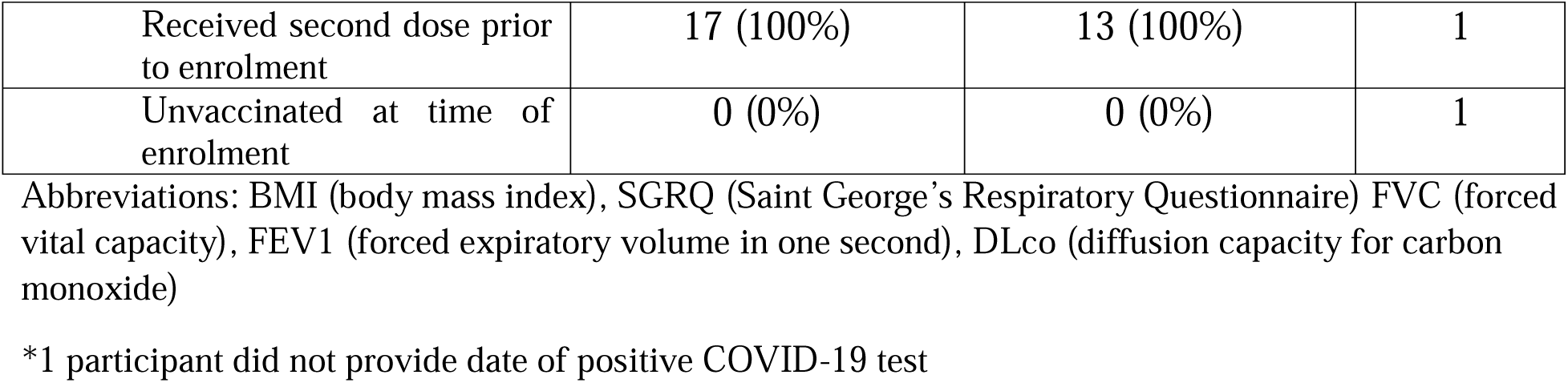
Demographics of participants with peripheral blood mononuclear cells. All values are expressed as mean ± standard deviation unless otherwise specified.

|  | Pulmonary long COVID (n=17) | Control (n=13) | P-value |
| --- | --- | --- | --- |
| Age, years | 44.5 ± 13.2 | 45.1 ± 17.4 | 0.9 |
| Male, n (%) | 6 (35%) | 6 (46%) | 0.6 |
| BMI, kg/m<sup>2</sup> | 25.8 ± 4.5 | 29.1 ± 6.0 | 0.2 |
| Tobacco smokers, n (%) |  |  | 0.7 |
| Current | 1 (5.9%) | 0 (0%) |  |
| Former | 4 (23.5%) | 3 (23.1%) |  |
| Never | 13 (70.6%) | 10 (76.9%) |  |
| Cannabis smokers (joint) , n (%) |  |  | 0.4 |
| Current | 1 (5.9%) | 2 (15.4%) |  |
| Never | 16 (94.1%) | 11 (84.6%) |  |
| Electronic Cigarettes, n (%) |  |  | 0.9 |
| Current | 1 (5.9%) | 1 (7.7%) |  |
| Never | 16 (94.1%) | 12 (92.3%) |  |
| SGRQ, total | 48.1 ± 16.8 | 8.5 ± 8.2 | 1.5 $10^{-7}$ |
| Symptoms | 57.9 ± 17.7 | 9.2 ± 7.6 | 8.3 $10^{-6}$ |
| Activity | 65.4 ± 21.3 | 16.7 ± 19.2 | 5.7 $10^{-5}$ |
| Impact | 35.1 ± 19.4 | 3.1 ± 4.5 | 1.6 $10^{-5}$ |
| Pulmonary Function Test |  |  |  |
| FVC, L | 4.1 ± 0.9 | 4.4 ± 1.1 | 0.5 |
| FVC, % of predicted | 106 ± 21 | 116 ± 14 | 0.12 |
| FEV<sub>1</sub>, L | 3.1 ± 0.56 | 3.5 ± 0.83 | 0.3 |
| FEV<sub>1</sub>, % of predicted | 100 ± 19 | 112 ± 15 | 0.1 |
| FEV<sub>1</sub>/FVC | 77.7 ± 8.4 | 79.6 ± 7.1 | 0.7 |
| DLco, % of predicted | 100 ± 17 | 111 ± 19 | 0.1 |
| Blood cell counts |  |  |  |
| White blood cells (x 10<sup>9</sup>/L) | 6.0 ± 1.7 | 5.9 ± 1.0 | 0.7 |
| Neutrophils (%) | 59.2 ± 9.6 | 57.8 ± 8.4 | 0.5 |
| Lymphocytes (%) | 31.3 ± 9.5 | 31.0 ± 6.5 | 0.8 |
| Monocytes (%) | 5.6 ± 1.3 | 5.4 ± 1.0 | 0.8 |
| Eosinophils (%) | 1.7 ± 0.82 | 2.7 ± 1.7 | 0.2 |
| COVID-19 Severity, n (%) |  |  | 0.04 |
| Hospitalized | 4 (23.5%) | 0 (0%) |  |
| Mild | 13 (76.5%) | 11 (84.6%) |  |
| Asymptomatic | 0 | 1 (7.7%) |  |
| Unexposed | NA | 1 (7.7%) |  |
| Vaccination status, n (%) |  |  |  |
| Vaccinated prior to infection | 6 (38%) (n=16)* | 5 (42%) (n=12) | 0.85 |
| Received first dose prior to enrolment | 17 (100%) | 13 (100%) | 1 |
| Received second dose prior to enrolment | 17 (100%) | 13 (100%) | 1 |
| Unvaccinated at time of enrolment | 0 (0%) | 0 (0%) | 1 |

Abbreviations: BMI (body mass index), SGRQ (Saint George's Respiratory Questionnaire) FVC (forced vital capacity), FEV1 (forced expiratory volume in one second), DLco (diffusion capacity for carbon monoxide)
\*1 participant did not provide date of positive COVID-19 test

### CyTOF of peripheral blood mononuclear cells and serum auto-antibodies

The PBMC samples were subjected to CyTOF. After quality control, 8,467,096 cells from these samples were used for downstream analyses, including clustering and annotation. A UMAP representation of 1,000,000 cells from these experiments is depicted in Figure 5A. Briefly, we generated 20 cell clusters, which included CD4+ T-cells, CD8+ T-cells, natural killer cells, B-cells, classical monocytes, non-classical monocytes, classical DCs, plasmacytoid DCs and stem cells. CD4+ T-cells were the most abundant cell type across all participants. PLC patients had a higher proportion of αβ T-cells expressing both CD8 and CD4 (double positive T-cells, p=0.0154), and a lower proportion of plasmacytoid DCs (p=0.027; Figure 5B). Absolute counts did not show any differences other than a decrease in plasmacytoid dendritic cells in the PLC group (p=0.048) (Supplemental Figure 7). None of the 38 marker proteins showed significant differential expression between PLC and control groups. However, CD8+ γδ T-cells and natural killer T-cells showed a decrease in CD138 protein expression in PLC participants according to the pseudobulk analysis (FDR=0.0112, FDR=0.0282, respectively). When stratifying by tertiles using SGRQ score, no significant trend emerged (Supplemental Figure 8).

**Figure 5.**
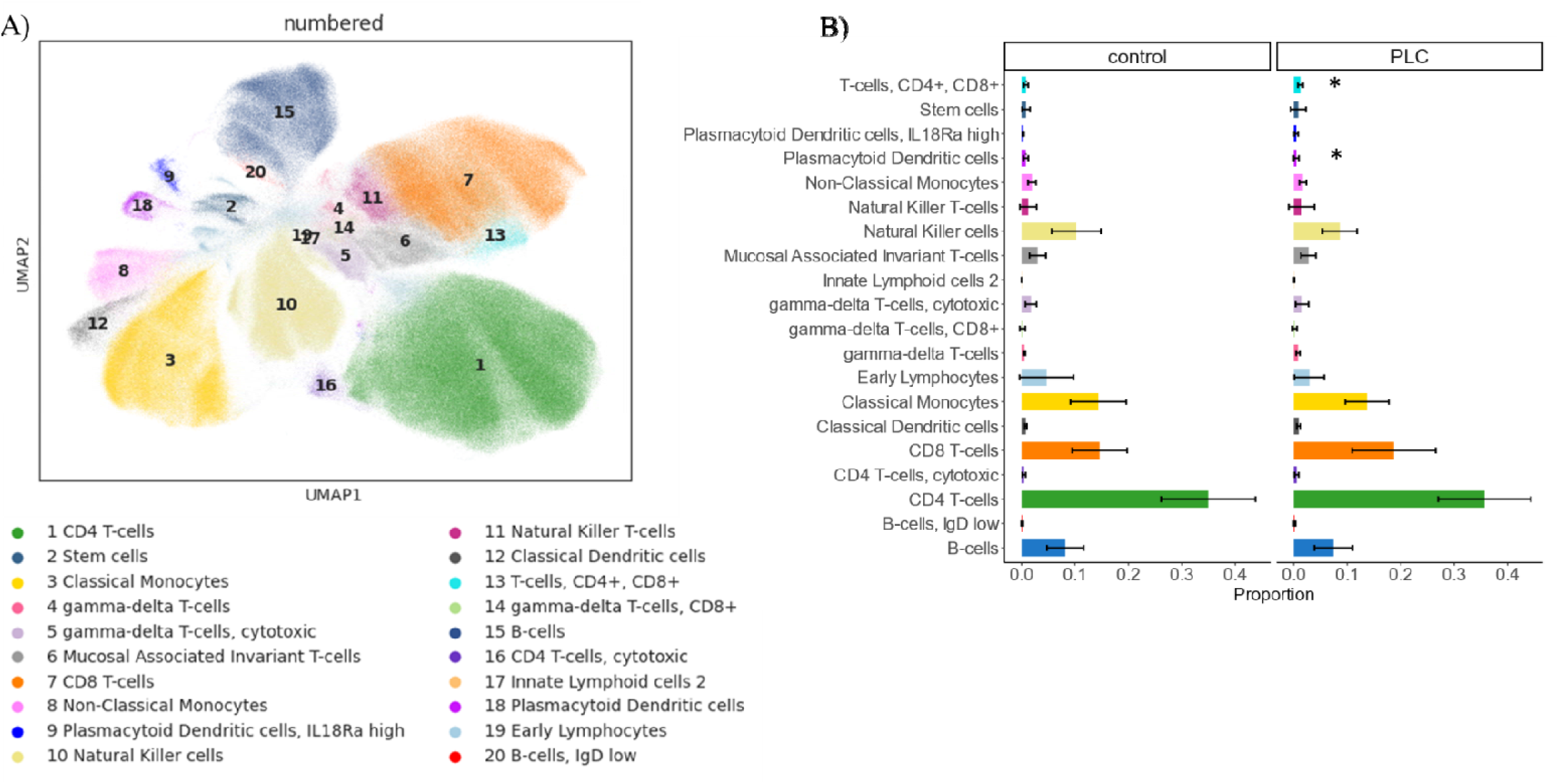
**A)** Uniform manifold approximation and projection for dimension reduction (UMAP) showing 1,000,000 PBMCs which underwent mass cytometry from pulmonary long COVID and control participants. Each dot represents a cell, and each cluster (denoted by number and colour) represents a cell population, grouped together based on protein expression. **B)** Composition plot showing the average cell proportions in control (left) and pulmonary long COVID (PLC, right) participants. T-cells CD8+, CD4+ (bright blue) were significantly increased in pulmonary long COVID participants while plasmacytoid dendritic cells (dark purple) were significantly decreased.

Of the 40 participant serum samples, which underwent the IgG autoantibody assay, 22 had PLC and the remaining 18 were controls. Further demographic information can be found in Supplemental Table 5. We measured 120 auto-IgG antibodies in serum and found that three of these were significantly increased in PLC (p<0.05) (Supplemental Figure 9). These were antibodies against genomic DNA (p=0.026), collagen II (p=0.048), and transcription intermediary factor gamma (TIF-γ, p=0.0049). Serum SARS-CoV-2 N protein concentrations did not significantly differ between groups (p=0.76).

## Discussion

Here, by applying CyTOF and scRNAseq technologies to well-phenotyped patient samples, we showed that persons with PLC have distinct changes in their immune landscape impacting both circulating and airway immune cells. Key finding includes an augmented cellular immune response in PLC, as evidenced by increased proportions of T-cell subsets in both PBMCs and BAL samples. This immune activation coincided with elevated serum autoantibodies in participants with PLC. In the BAL fluid of PLC participants, transcriptomic alterations were identified in T-cells, DCs, and alveolar macrophages, consistent with increased anti-viral, Th1 immune responses. Remarkably, these changes were observed in samples collected many months after their acute infection and there was no evidence of viral persistence in the airways. Taken together, these data suggest that pulmonary symptoms in long COVID may be driven by persistent inflammation in the distal lung related to increased cellular (T-cell-mediated) immunity, possibly associated with autoimmunity.

Our results are consistent with previous studies. For example, one previous study found heightened T-cell responses in patients with radiological abnormalities three months post-hospital discharge^35^. Our findings show that this T-cell mediated response is present even many months (and years) post-infection. Elevated levels of inflammatory mediators in the blood, including interferons, were detected in patients experiencing persistent fatigue, dyspnea, or chest pain^6^. Additionally, home-recovered patients with long COVID exhibited a decrease in the number of effector memory T-cells in blood, which may indicate that they have been recruited to other tissues (such as the lungs, as observed here)^36^. A murine study demonstrated that the interactions between CD8+ T-cells and macrophages resulted in increased IL-1β, which may drive ongoing airway inflammation^37^. Disruption in other cell types, such as neutrophils, have also been reported in PLC lungs^12^. Neutrophils have been described to play a major role as effector cells in many autoimmune diseases^38^. Other evidence for autoimmunity as a driver for PLC includes a study by Son et. al. which found elevated serum auto-antibodies in patients with long COVID^39^.

While long COVID is among the more popular post-acute viral sequelae, other post-infection conditions have been reported in the literature. For instance, mosquito-borne flavivirus infections damage cells leading to long-term neurologic changes^40^. Both SARS-CoV-2 and influenza have also been shown to alter gene expression in hamster spinal cords^41^. In these hamsters, interferon pathways were more common during the acute stage and after 31 days, metabolic pathways became more common^41^. Other SARS viruses, including SARS-1 and MERS, showed long term effects, including a decreased quality of life, mental health complications, and in some cases residual pulmonary lesions on CT, which were present even after a decade post-infection^42^. IFN-7 is shown to be upregulated in CD8+ T-cells as a means to recruit effector cells which maty lead to lung function impairment following influenza-infection^43^. In elderly patients especially, influenza is known to have an inflammatory sequela, resulting in scar tissue in the parenchyma and PPAR-y deficient myeloid cells which result in more collagen deposition in the parenchyma^44^. Pro-fibrotic alveolar macrophages are also present in the lung tissue of patients with idiopathic pulmonary fibrosis (IPF)^45,46^. Lung-resident T-cells have a role in IPF as well. CD8+ T-cells in the parenchyma is associated with decreased lung function^47^. Activated CD4+ T-cells in BAL are higher in patients with asymptomatic ILD compared to healthy participants^47^. In our populations, all CD4+ T-cells were upregulated in pulmonary long COVID participants, a highly symptomatic group. In PBMCs, IPF patients have an increase in classical monocytes and T-regulatory cells, compared to our findings in long COVID which shows an increase in plasmacytoid dendritic cells and CD8+, CD4+ T-cells^48^. In both cases, there is some evidence for the recruitment of immune cells from the blood into the lungs^48^.

Our study is among the first to apply scRNAseq to BAL samples to characterize the distal airways of long COVID patients with pulmonary symptoms. Previous studies using BAL samples primarily focused on acutely ill patients infected with SARS-COV-2^49^. In contrast, our cohort included individuals with mild acute episodes of COVID-19 but continued to have persistent pulmonary symptoms post-infection. Despite the severity of their symptoms, most patients had normal pulmonary function tests and CT scan results. We identified significant correlations between BAL CD4+ T-cell proportions and SGRQ total scores, a measure of health-related quality of life. Differential expression analysis revealed elevation of *MTRNR2L12*and *HLA-DRB5* in BAL samples of PLC patients. *MTRNR2L12* is known to be a negative regulator of apoptosis, and has been shown to be upregulated in BAL cells in acute COVID-19^50^. Genes associated with interferon responses, such as *IFIT*, *IFITM, and ISG15*, were also upregulated in PLC patients, reflecting sustained anti-viral responses in the airways^51,52^. Among these candidates, *IFITM*s are thought to serve as co-entry receptors for SARS-CoV-2^51^. Additionally, *CCL4*, a chemokine associated with type 1 immunity and negatively regulated by type 2 cytokines such as IL-4, was also upregulated in CD4+ T-cells^53,54^. In parallel, these findings were associated with an increase in circulating double-positive T-cells. We posit that these T-cells enter the airways, where they differentiate into CD4+ T-cells in response to an inflammatory insult such as those related to autoimmunity. Double-positive T-cells have been previously associated with autoimmunity, as they have been identified in the skin of those with atopic dermatitis and the joint fluid of patients with rheumatoid arthritis^55^. Furthermore, an increase of auto-antibodies against collagen II is consistent with findings that suggest a higher prevalence of autoimmune disease, such as rheumatoid arthritis, in COVID-19 patients^56^. Anti-genomic DNA is also a hallmark of autoimmunity and is increased in patients with systemic lupus erythematosus^57^. Finally, TIF-γ autoantibodies are associated with autoimmune conditions relating to the skin, most notably dermatomyositis^58^.

There are several limitations to the study. First, while we have characterized both lung and circulating immune cells, we lacked a functional model to assess mechanisms of pathogenesis. Second, we accessed blood and BAL samples at one time point, which limits causal inferences. Third, while we observed an increase in CD4+ T-cells in the BAL of PLC patients by scRNAseq and CyTOF, the high number of cells required for the latter assay reduced the overall sample size, which limited our statistical power. Despite this, we were able to fully characterize 105,836 cells by scRNAseq and 456,631 by CyTOF, making this one of the largest datasets for BAL, which is a relatively difficult sample to obtain. Fourth, our study recruited predominantly younger persons without significant co-morbidities or pre-existing lung disease. It is well established that long COVID is a heterogeneous disease^59^. As such our study may not be generalizable to older patients with multiple co-morbidities. Fifth, as most patients in the study had PLC for less than 2 years, the long-term trajectory of airway inflammation remains uncertain. Moreover, the time between the initial infection and bronchoscopy or blood draw was quite variable among participants, though all participants were at least three months post-infection.

In summary, we found that PLC patients have an altered immune landscape indicating persistent small airway inflammation, possibly related to perturbed adaptive immune responses and autoimmunity. These findings highlight T-cells as possible therapeutic targets for long COVID patients with persistent pulmonary symptoms. Future studies should address these limitations by incorporating longitudinal analyses and functional models to elucidate mechanisms and refine therapeutic approaches.

## Data Availability

All data produced in the present study are available upon reasonable request to the authors

## Acknowledgements

This work was made possible with the support of the staff from the Biomedical Research Centre at the University of British Columbia (UBC), including Yvonne Chung, Stephen Yu, and Bernie Zhao for their technical assistance with single-cell RNA sequencing. We would also like to extend our gratitude to Andrew Warkman and Adrianna Suarez from 10X genomics, for their guidance on the sequencing workflow and bioinformatic analysis. Special thanks to Mark Hamer from the UBC’s CyTOF core for his technical support with CyTOF, Micheal Williams from the UBC Antibody – Protein Engineering – Biologics core for assistance with antibody procurement and conjugation, Laura Burns from Providence Healthcare for coordinating and running the Roche antibody assay, and Indu Raman from the University of SouthWestern Texas Microarray core for coordinating and assisting with the autoantibody IgG assays.

## Supplemental Materials

### Methods

#### Participant Recruitment

Through advertisements in Vancouver, Canada and the surrounding region, we recruited persons who had been previously infected with SARS-CoV-2 and were clinically stable for at least 3 months post-infection. We excluded persons who had received a solid organ transplant or were taking an anti-coagulant or systemic immunosuppressive therapies. Patients who were unable to undergo bronchoscopy were excluded from the bronchoscopy cohort. Additionally, participants underwent pulmonary function testing (PFTs), low-dose computerized tomography (CT) as previously described^11^. Acute infection severity was divided based on whether participants were hospitalized, home-recovered (i.e. mild) or asymptomatic.

#### Bronchoalveolar Lavage acquisition

After participants were lightly sedated, a fiberoptic bronchoscope (Olympus Corporation, Tokyo, Japan) was orally inserted and passed into the lower airways. The bronchoscope was wedged carefully into a subsegmental bronchus in the right middle lobe (or lingula if the right middle lobe was not accessible) and a bronchoalveolar lavage (BAL) procedure was performed by instilling warm sterile saline until a return volume of 30 mL or a total instilled volume of 200 mL was reached. An aliquot of the recovered BAL was first processed for cell count using an automated counter.

#### Blood Collection

Blood was drawn from participants during their first visit. A blood tube containing EDTA was used to collect six mLs of blood. The blood was then run on the Advia 2120 (Siemens, Munich, Germany) to obtain a complete blood count and differentials. Peripheral blood mononuclear cells (PBMCs) were isolated using the SepMate™ PBMC Isolation method developed by STEMCELL Technologies (Vancouver, Canada). Once isolated, the cells were cryopreserved in fetal bovine serum (FBS) with 10% dimethyl sulfoxide (DMSO). The PBMCs were then frozen down in a Mr. Frosty™ Freezing Container (Thermo Fisher Scientific, Waltham, USA) with isopropyl alcohol at -80°C for a minimum of two hours before being transferred to liquid nitrogen. Serum was also collected as this time and aliquoted into a maximum of five aliquots of 250 μL. PBMCs were collected and cryopreserved for all participants who attended their first visit after April 1^st^, 2022 until January 2024.

##### Serum Antibody Assays

The Elecsys® anti-SARS-CoV-2 by Roche Diagnostics (Indianapolis, USA) was used to confirm the presence of anti-N antibodies in participants who reported previously existing SARS-CoV-2 infections, as previously described^60^. The serum of all participants whose blood we collected underwent this assay.

Analysis of the autoantibody microarray as performed using R. A signal to noise ratio of greater than three was considered a true signal, and the normalized signal intensity between control and PLC participants was compared using Wilcoxon’s rank sum test. P-values were adjusted to false-discovery rates (FDR) to adjust for multiple-comparisons. Forty participants were selected for this assay, including all participants who had PBMCs isolated from CyTOF as well as all participants who underwent bronchoscopy.

#### Bronchoalveolar Lavage processing

After acquisition of the BAL from right middle lobe or lingular segment, an aliquot of the recovered sample was first processed for cell count using an automated counter. For single-cell RNA-sequencing processing, the remaining BAL sample was then poured through a 70 µm strainer and spun at 400 g for 10 mins at 4 °C. The supernatant was removed, and the pellet was resuspended in a 10 mL solution of DPBS in 0.4% bovine serum albumin (BSA). The sample was spun again and resuspended in either 500 µL or 1000 µL depending on the pellet size. A cell count was then performed using Trypan blue in a hemocytometer.

Single Cell Suspensions were loaded onto the 10x Genomics Chromium X for capture in droplet emulsion. Libraries were prepared using the Chromium Next GEM Single Cell 3’ Kit v3.1 and the standard protocol was followed for all steps. Libraries were then sequenced on an Illumina NextSeq 2000. 10x Genomics Cell Ranger 6.0 was used to perform Demultiplexing, Alignment, Counting, Clustering, and Differential expression.

If the BAL had over one million cells leftover, we isolated a minimum of 700,000 cells and spun down the pellet at 300 g for five minutes. The supernatant was removed and replaced with an FBS + 10% DMSO solution. They were then frozen using the same protocol as the PBMCs for CyTOF analysis.

The remaining cells were processed for staining as described by Ho et. al^61^. The BAL fluid was strained, spun, and resuspended. A portion of the sample was stored in QIAzol (Qiagen, Cat. #79306) and used for bulk RNA-sequencing. The supernatant was kept and concentrated using Pierce™ Protein Concentrators, 3K Molecular Weight Cut-Off (ThermoFisher Scientific, Cat. #88515) by spinning 5-6 mL of the supernatant for 90 mins at 25°C at 2500 g.

Sample quality control was performed using the Agilent 2100 Bioanalyzer or the Agilent 4200 Tapestation. Qualifying samples were then prepped following the standard protocol for the Illumina Stranded mRNA prep (Illumina). Sequencing was performed on the Illumina NextSeq2000 with Paired End 59bp × 59bp reads. Sequencing data was demultiplexed using Illumina’s BCL Convert. De-multiplexed read sequences were then aligned to the Homo sapiens (hg38 no Alts, with decoys) reference sequence using DRAGEN RNA app on Basespace Sequence Hub.

The reads of the RNA sequencing files were then mapped to the SARS-CoV-2 viral transcriptome using salmon version 0.13.1^62^.

#### Anti-N SARS-CoV-2 ELISA

The serum and concentrated BAL supernatant were then used to run an enzyme-linked immunosorbent assay (ELISA) to measure human SARS-CoV-2 Nucleocapsid. The samples were run as directed on two plates developed by AFG BioScience (product code: EKCOV010). Sample concentrations were then normalized based on total recovered volume of BAL.

#### Cytokine measurements

Cytokines in serum and concentrated BAL supernatant were then plated on a U-Plex multi-plex cytokine assay (Meso Scale Diagnostics, Maryland, USA). Analytes measured include GM-CSF, IFN-γ, IL-1β, IL-4, IL-5, IL-6, IL-8, IL-10, IL-12p70, TNF-α, IFN-α2a, IFN-β, IL-1Rα, IL-7, IL29/IFN-λ1, MIP-1α, MIP-1β, and IL-9.

#### Cytometry by time-of-flight

The cells were thawed and immediately quenched in RPMI 1640 media with 10% FBS and 25U nuclease (Thermo Fisher Scientific, Cat #88700). The samples were then washed twice in this solution before being incubated with a Cell-ID™ Intercalator-Rh (Fluidigm, Cat. #201103A). Following the incubation, the cells were washed before an Fc blocker was applied. After fifteen minutes, the samples were washed again and stained with an antibody cocktail (Supplementary Materials Table 1 and 2). The cells were washed again with MaxPar® MCSB (Fluidigm, Cat. #201068) and fixed with Cell-ID™ Intercalator-IR (Fluidigm, Cat. #201192A). The following day, the cells are washed in MilliQ water and calibration beads were added.

#### Data Analysis

##### CyTOF

Following normalization, events were manually gated using the FlowJo gating software (BD Biosciences) to exclude dead cells, debris, beads, and non-nucleated cells. A batch control was run for each of the three runs of the PBMCs to account for batch effect.

##### Single-cell sequencing

Quality control, (including the removal of dead cells via mitochondrial gene expression count (threshold of 15%) and doublet removal using the Scrublet package), normalization, batch correction using the batch-balance k-nearest neighbor (BBKNN) algorithm, dimensionality reduction (notably PCA and UMAP), and clustering (using the Leiden algorithm) was performed on the 10X Matrix files. Before clustering, mitochondrial, ribosomal, hemoglobin and MALAT-1 genes were removed. Any epithelial cells were considered contamination and were removed for downstream analysis. Pseudobulk analysis was applied for differential gene expression analysis. Briefly, pseudobulk is a bioinformatic technique which adds the gene expression of all cells from one participant to generate one gene expression profile for each cell type^63^. This allows for more robust, less outlier-prone gene expression analysis. The SC-CODA model uses one cell type as a reference and evaluates the other cell types based on the assumption that the reference is unchanged between the groups. In our analysis, we sequentially selected each cell type as the reference and considered cell types that were increased more than 50% of the time to be statistically credible.

## Author Contributions

EG and DDS prepared the first draft of the manuscript. Acquisition of data was performed by EG, ICK, SC, FVG, JWY, MM, CG, DY, RLE, TS, CYC, JSWY, WY, and JML. EG, SS, XL analyzed the data.

Interpretation was done by EG, DDS, KMM, SS, ICK, SC, and HYP. All authors reviewed the manuscript and approved the final draft.

---

**Table 1.**
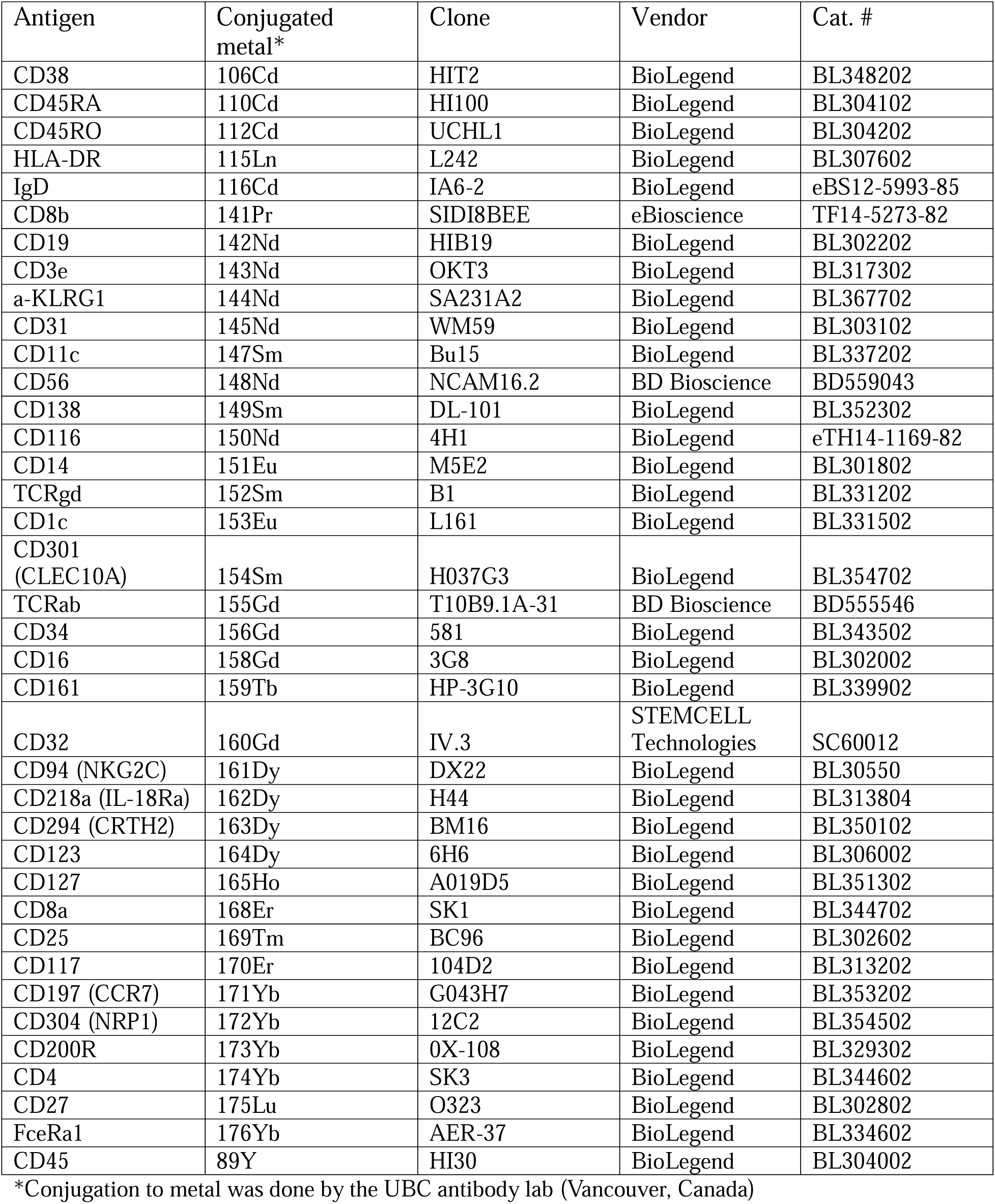
Peripheral blood mononuclear cells antibody panel.

**Table 2.**
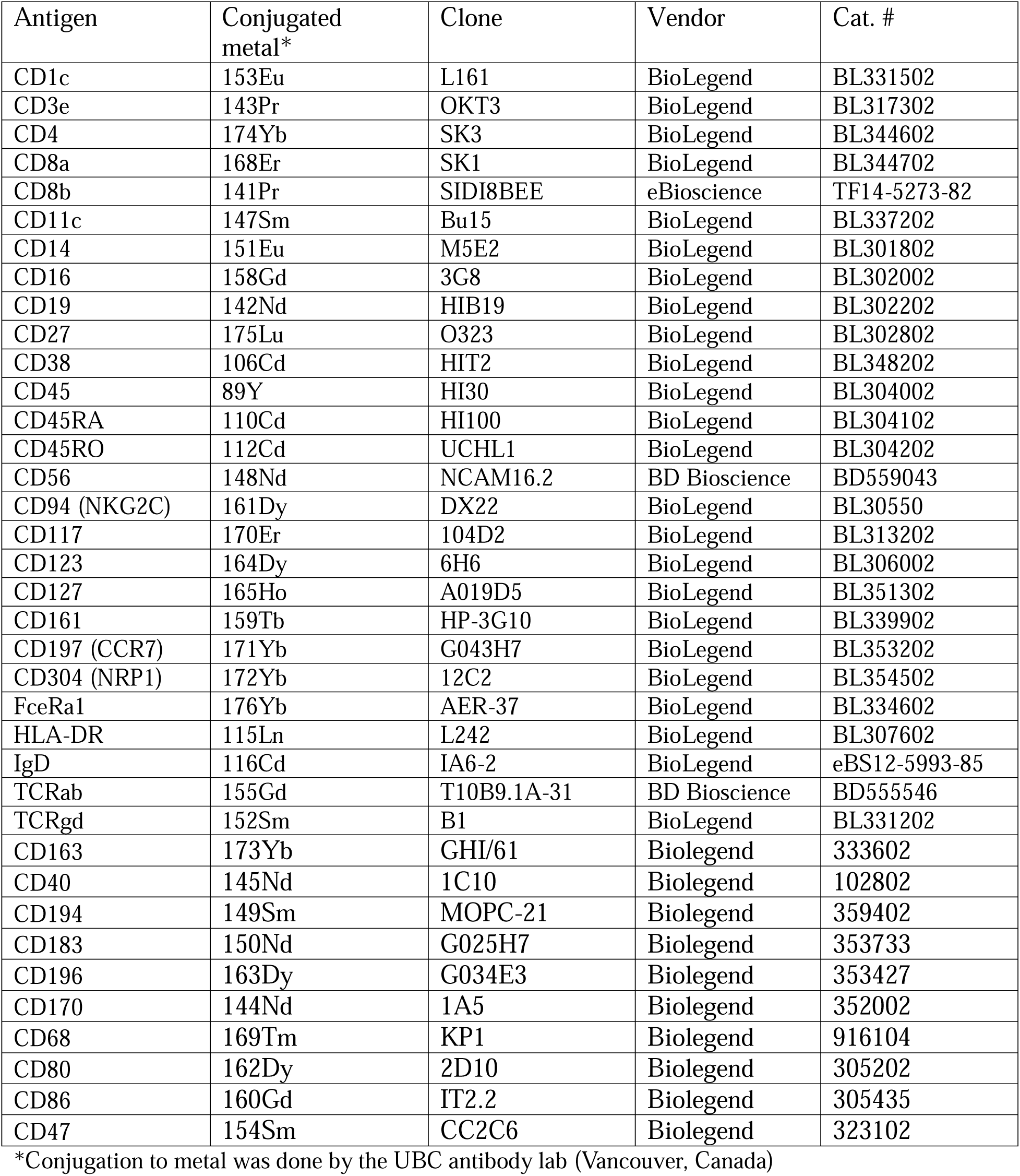
Bronchoalveolar lavage antibody panel.

**Table 3.**
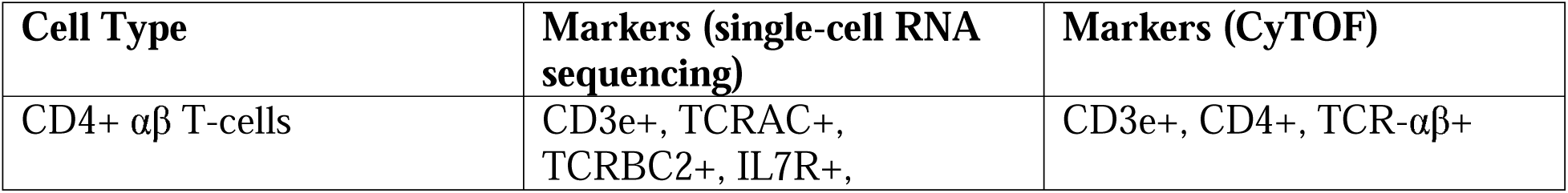
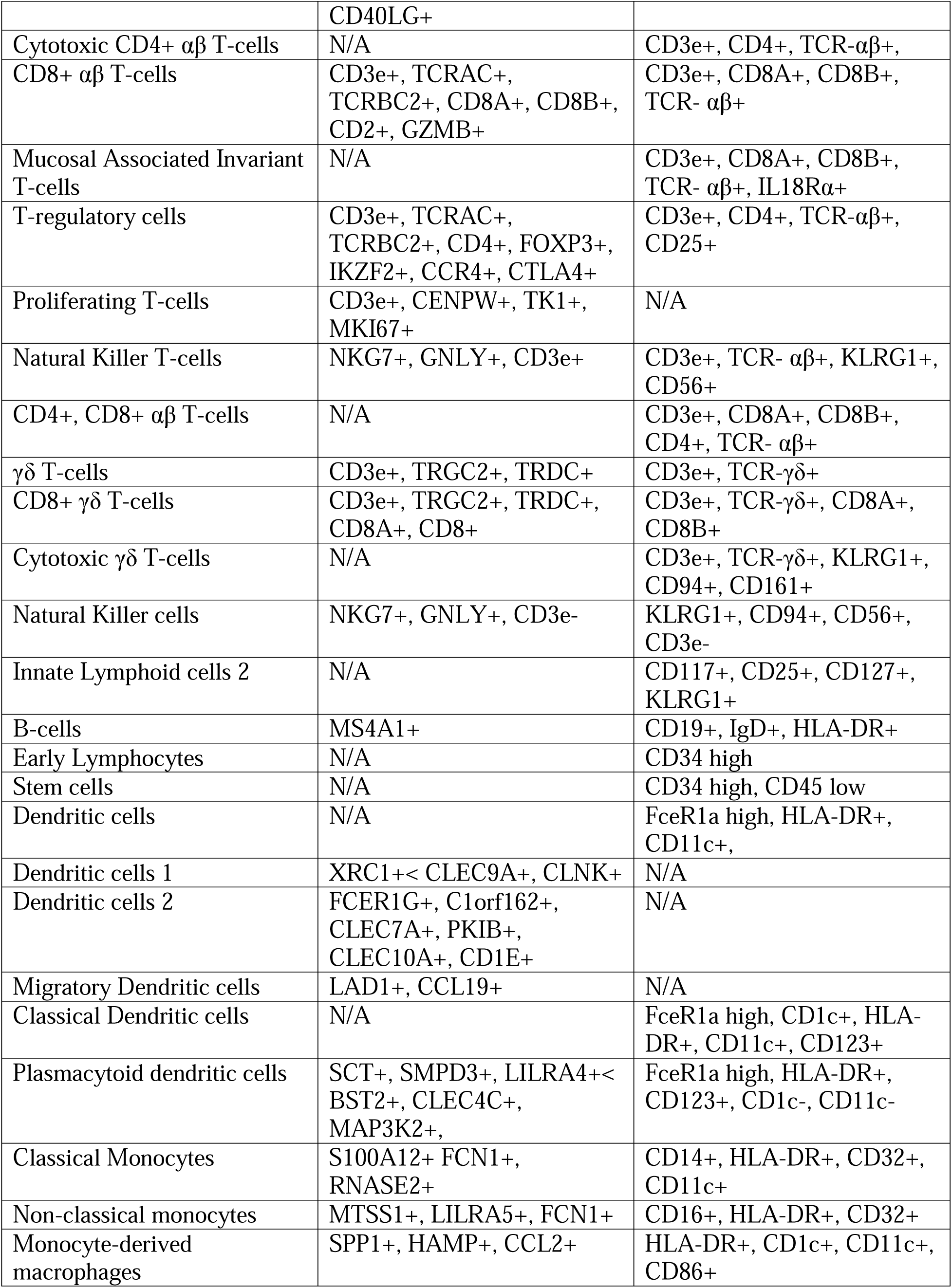
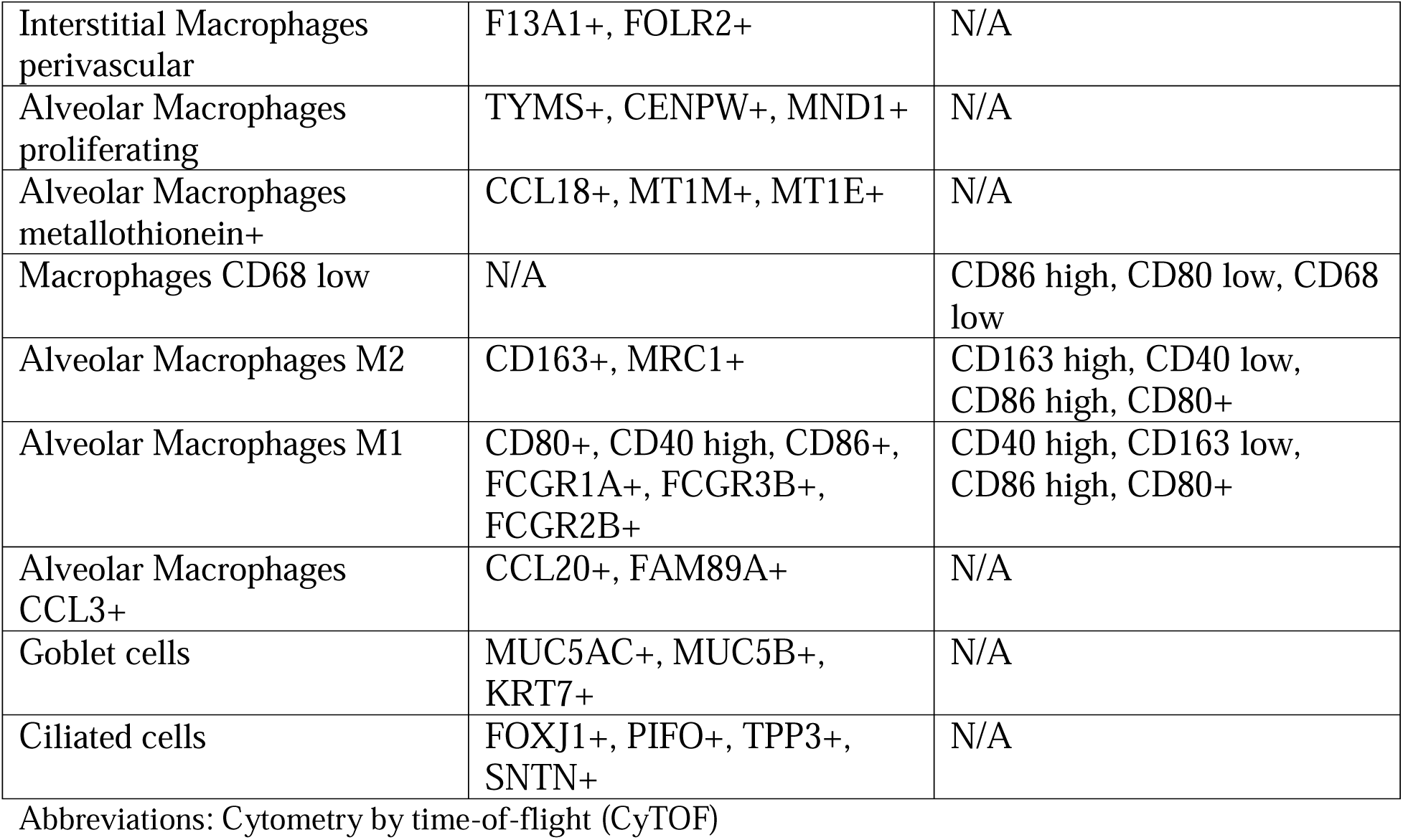
Annotation guide for single-cell RNA sequencing and cytometry by time-of-flight.

**Table 4.**
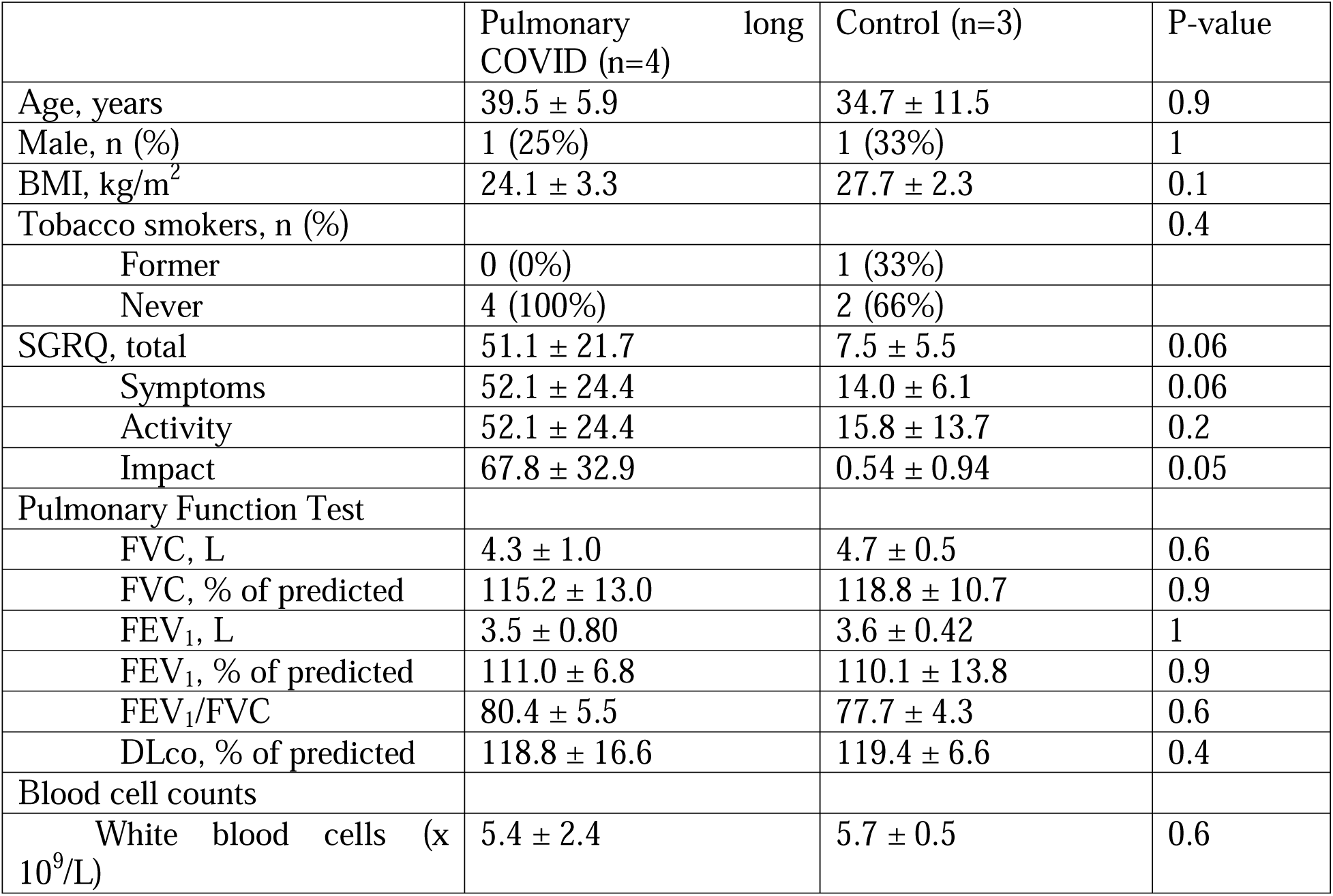
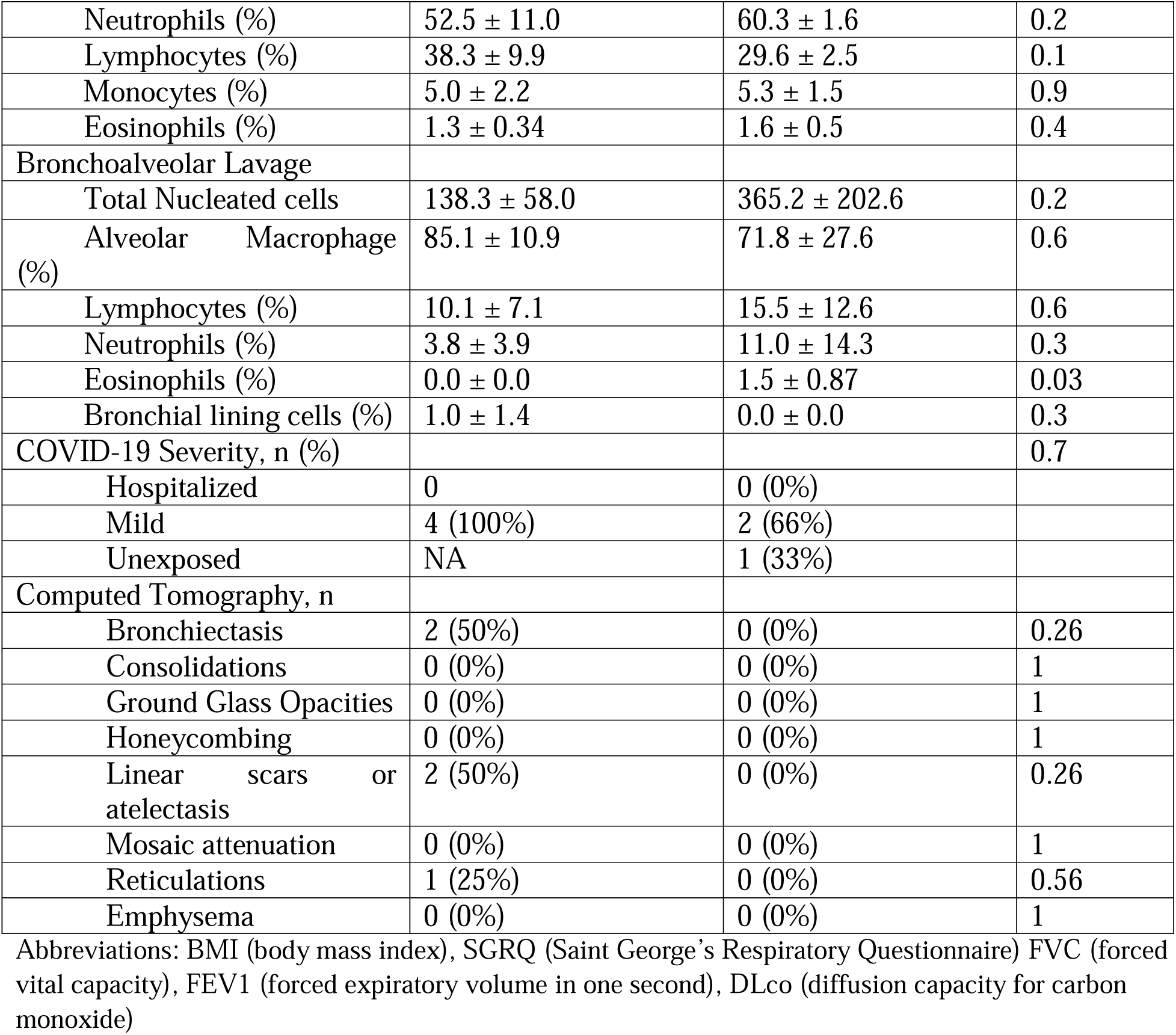
Demographics of participants with CyTOF data of bronchoalveolar lavage. All values are expressed as mean ± standard deviation unless otherwise specified.

**Table 5.**
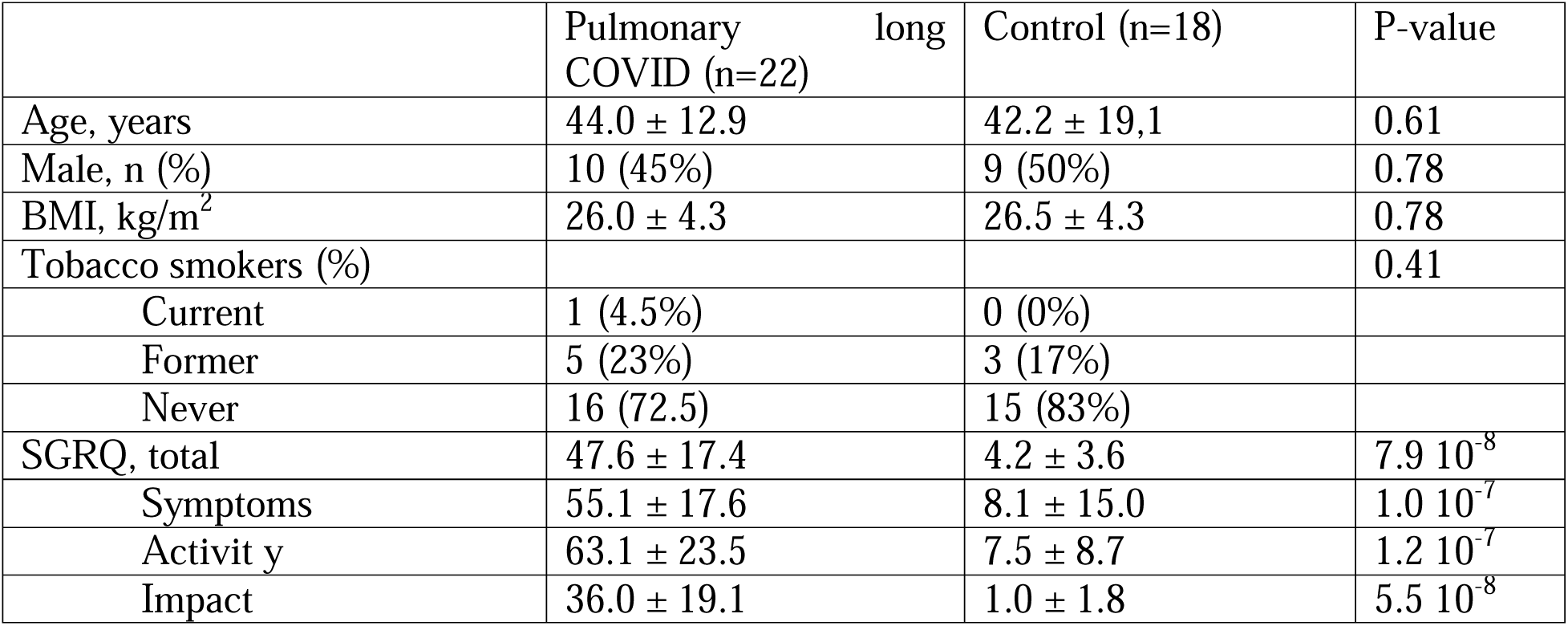
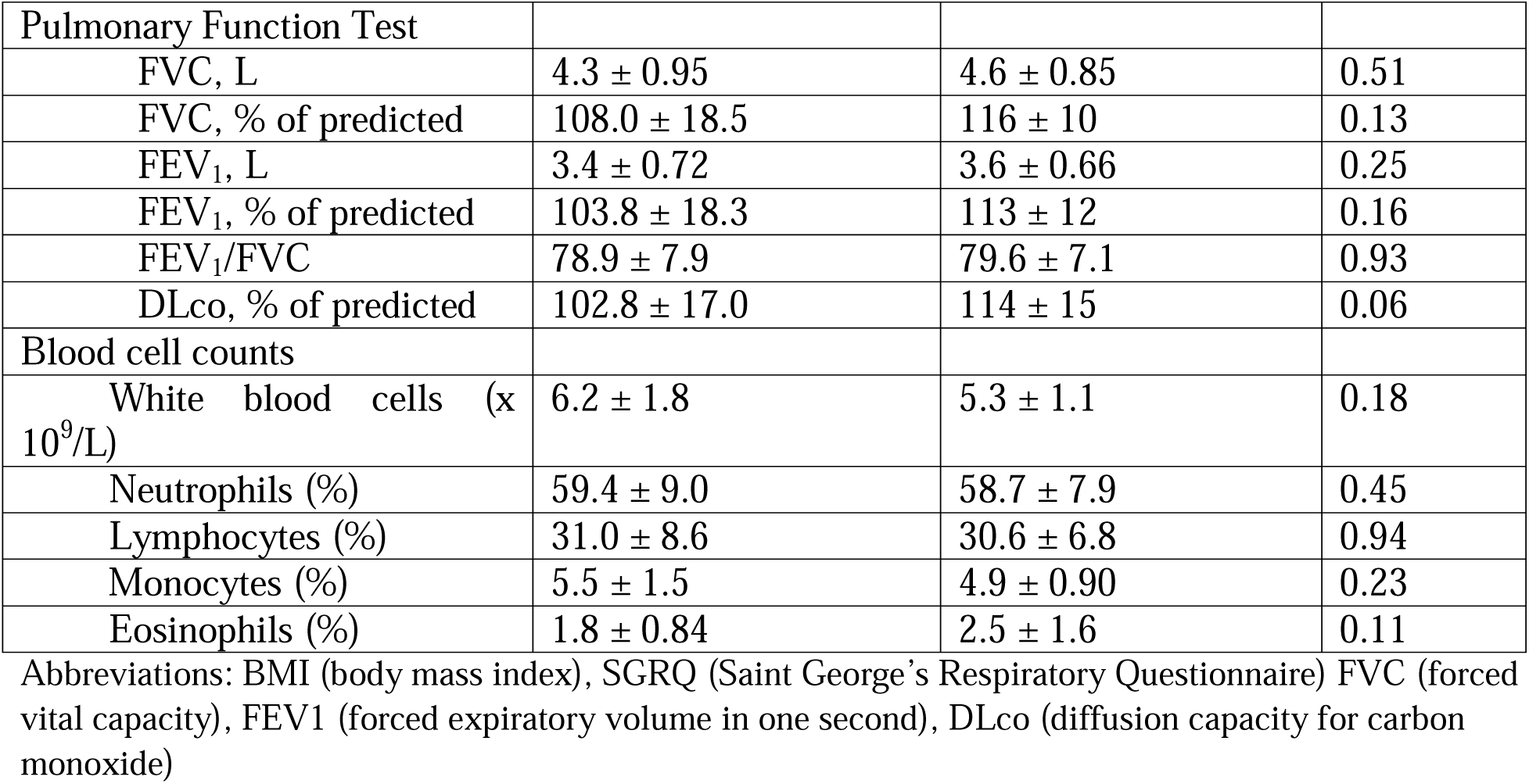
Demographics of participants with serum autoantibody data. All values are expressed as mean ± standard deviation unless otherwise specified.

**Supplemental Figure 1.**
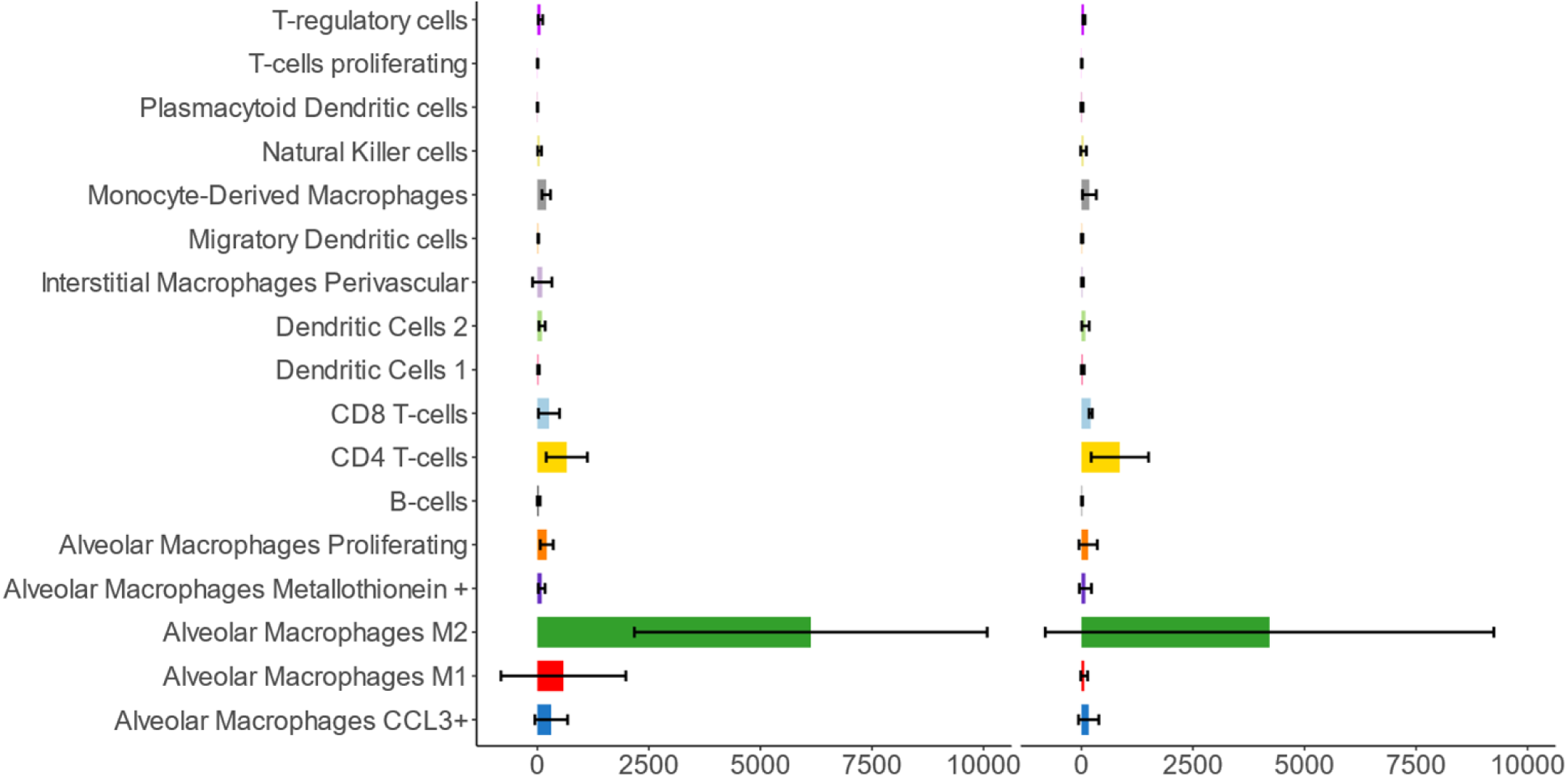
Composition plot showing the average absolute number of cells and standard deviation of the BAL single-cell RNA sequencing in control (left) and pulmonary long COVID (PLC, right) participants.

**Supplemental Figure 2.**
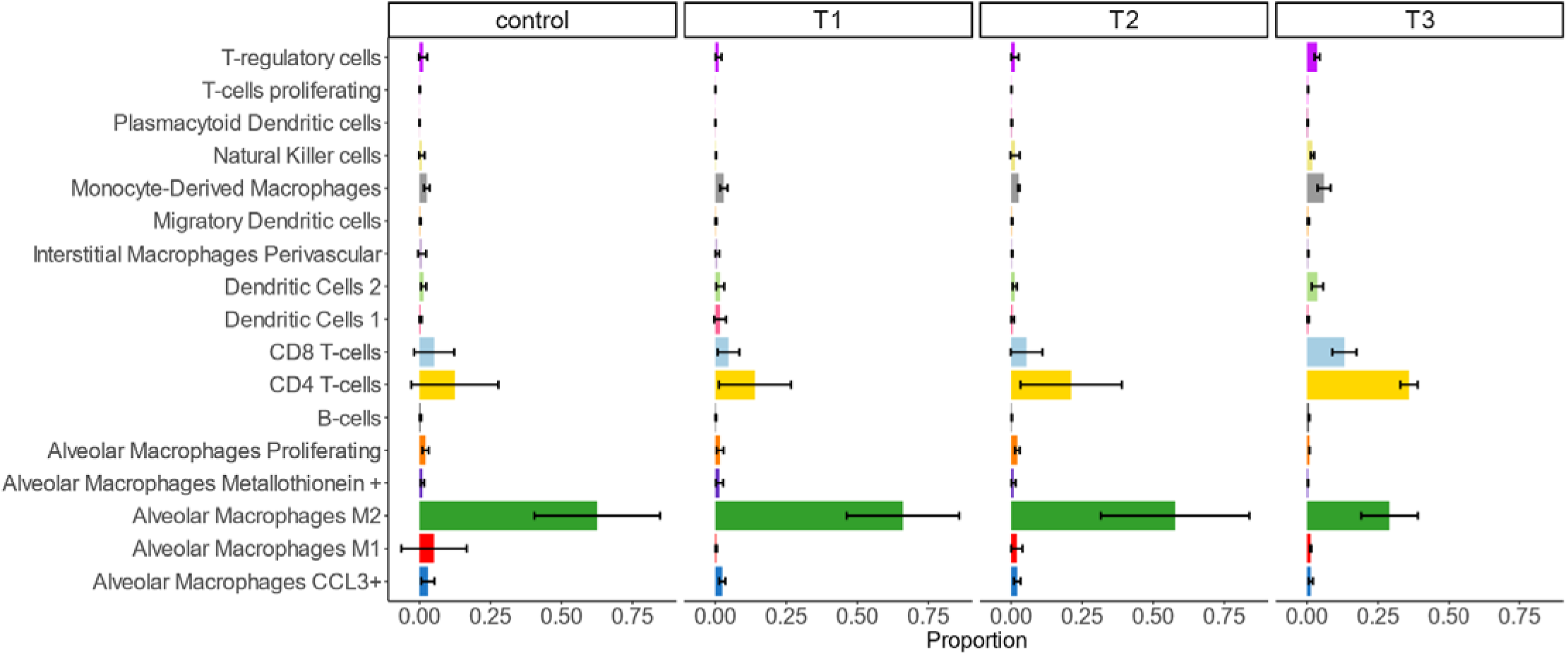
Composition plot of the average cell proportions and standard deviation of the BAL from single-cell RNA sequencing by SGRQ score. PLC participants were divided by tertile, where T3 had the highest SGRQ scores, and T1 the lowest. CD4+ T-cells increase with each tertile, while alveolar M2 macrophages decrease.

**Supplemental Figure 3.**
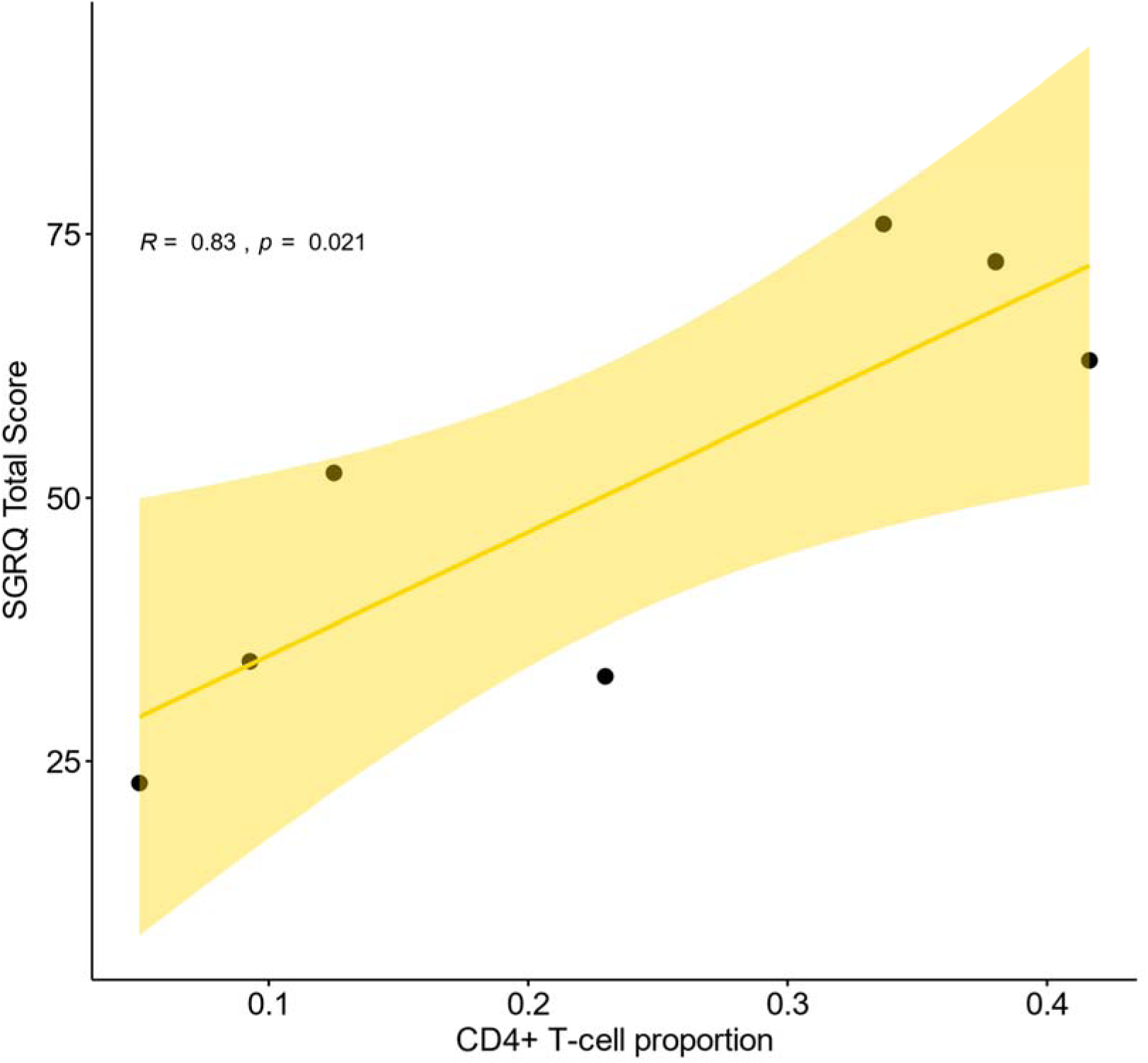
Spearman’s correlation between Saint George’s respiratory questionnaire’s total score and bronchoalveolar lavage CD4+ T-cells in pulmonary long COVID participants. The highlighted area represents the 95% confidence interval. Each point represents one participant. The relationship is considered strong, positive (R=0.83) and significant (p=0.021).

**Supplemental Figure 4.**
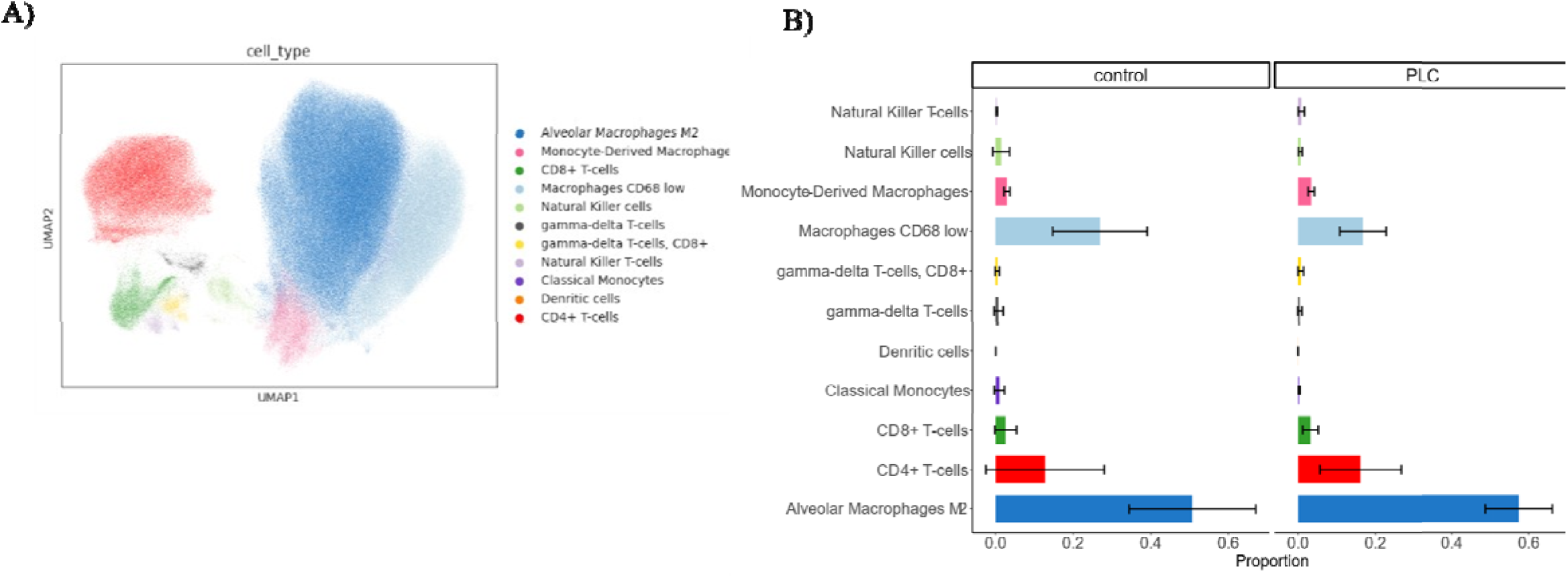
Results from CyTOF of the bronchoalveolar lavage, A) UMAP representation of all 456,631 cells from both PLC and control participants denoted by cluster (colour). B) Composition plot of the average cell proportions and standard deviation between PLC (right) and control (left). CD4+ T-cells were slightly increased in PLC participants but not significantly.

**Supplemental Figure 5.**
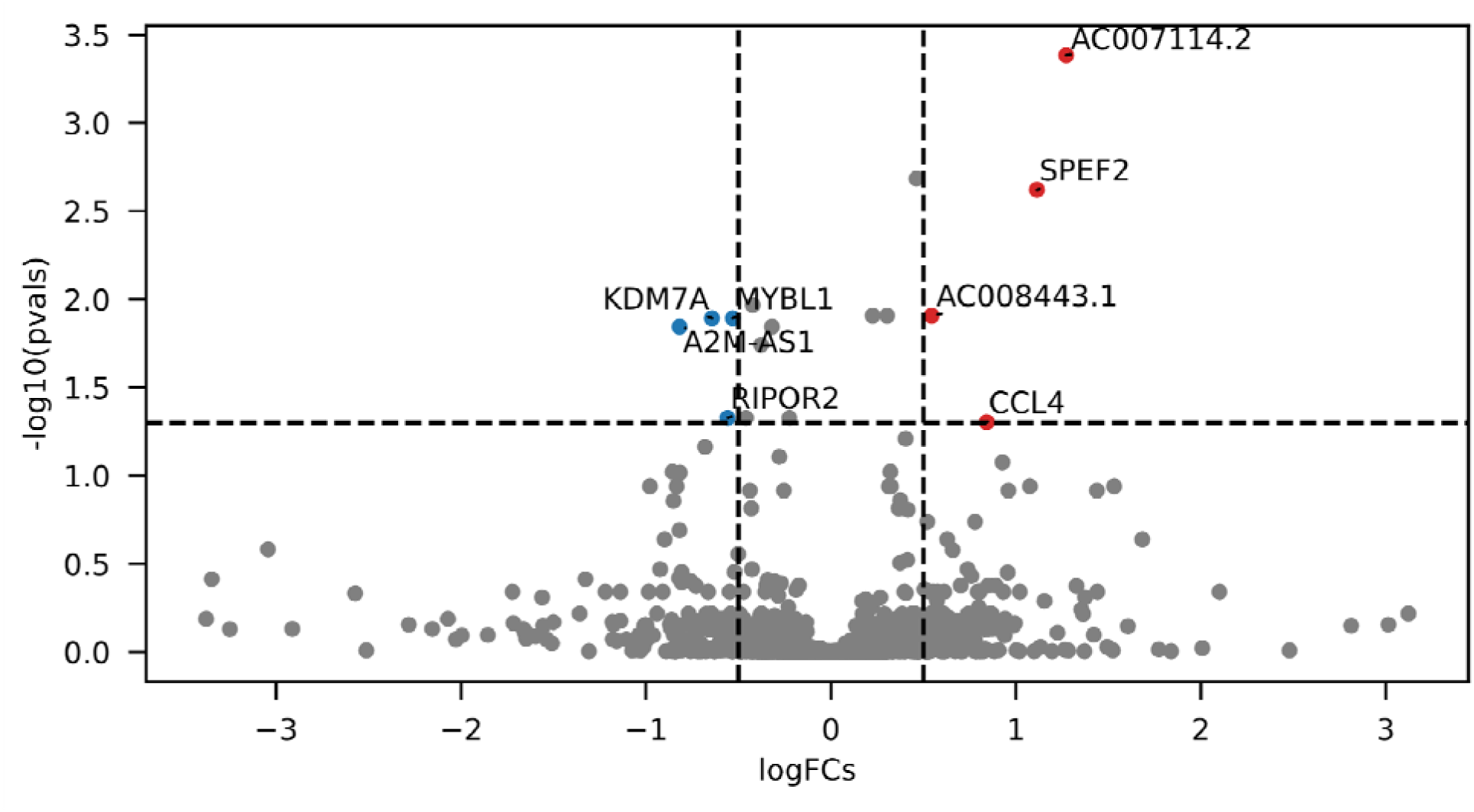
Volcano plot depicting differential expressed genes determined by pseudobulk analysis in CD4+ T-cells

**Supplemental Figure 6.**
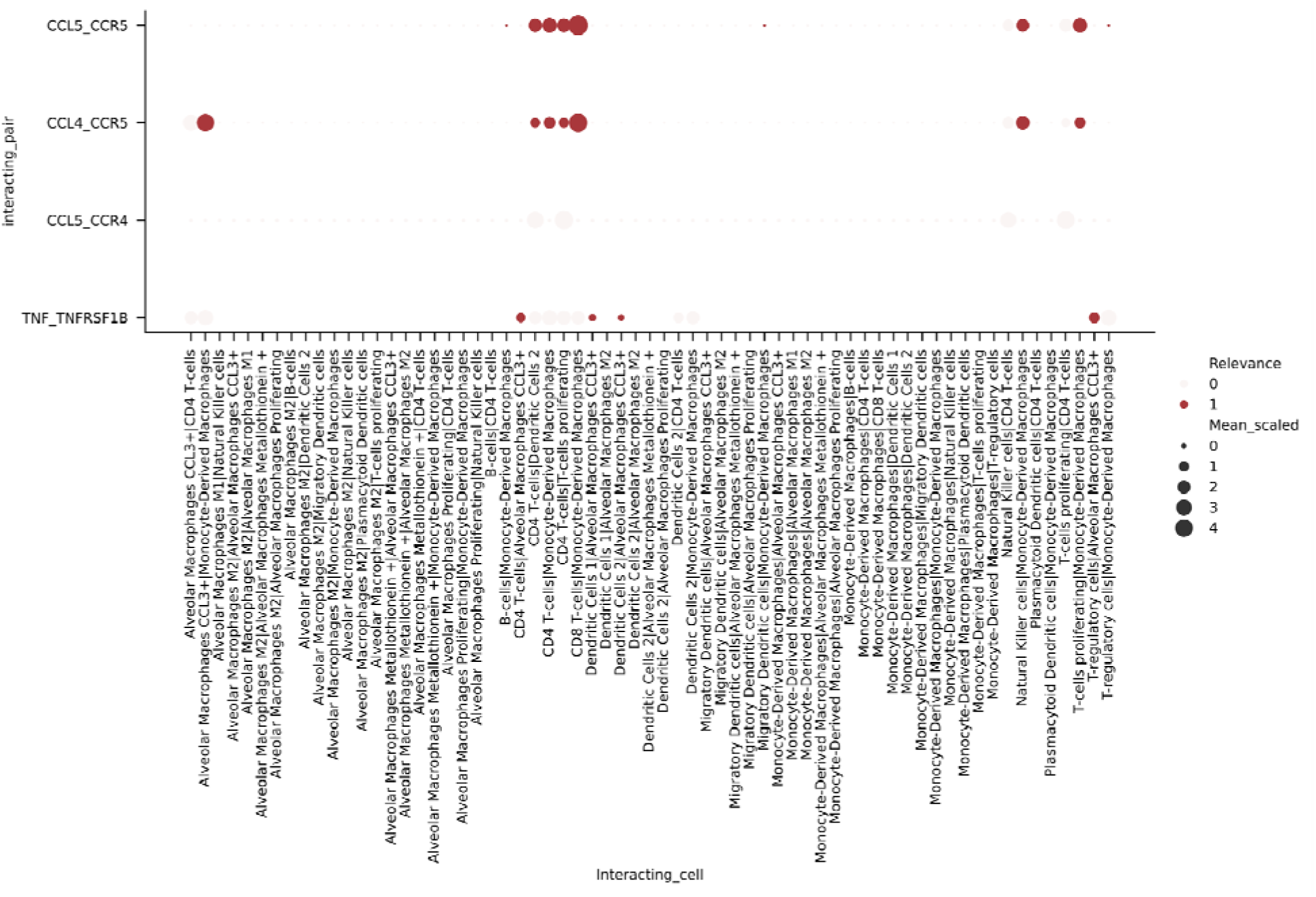
Relevant Interaction plot depicting important intercellular interactions using the single-cell RNA sequencing data of bronchoalveolar lavage in pulmonary long COVID participants.

**Supplemental Figure 7.**
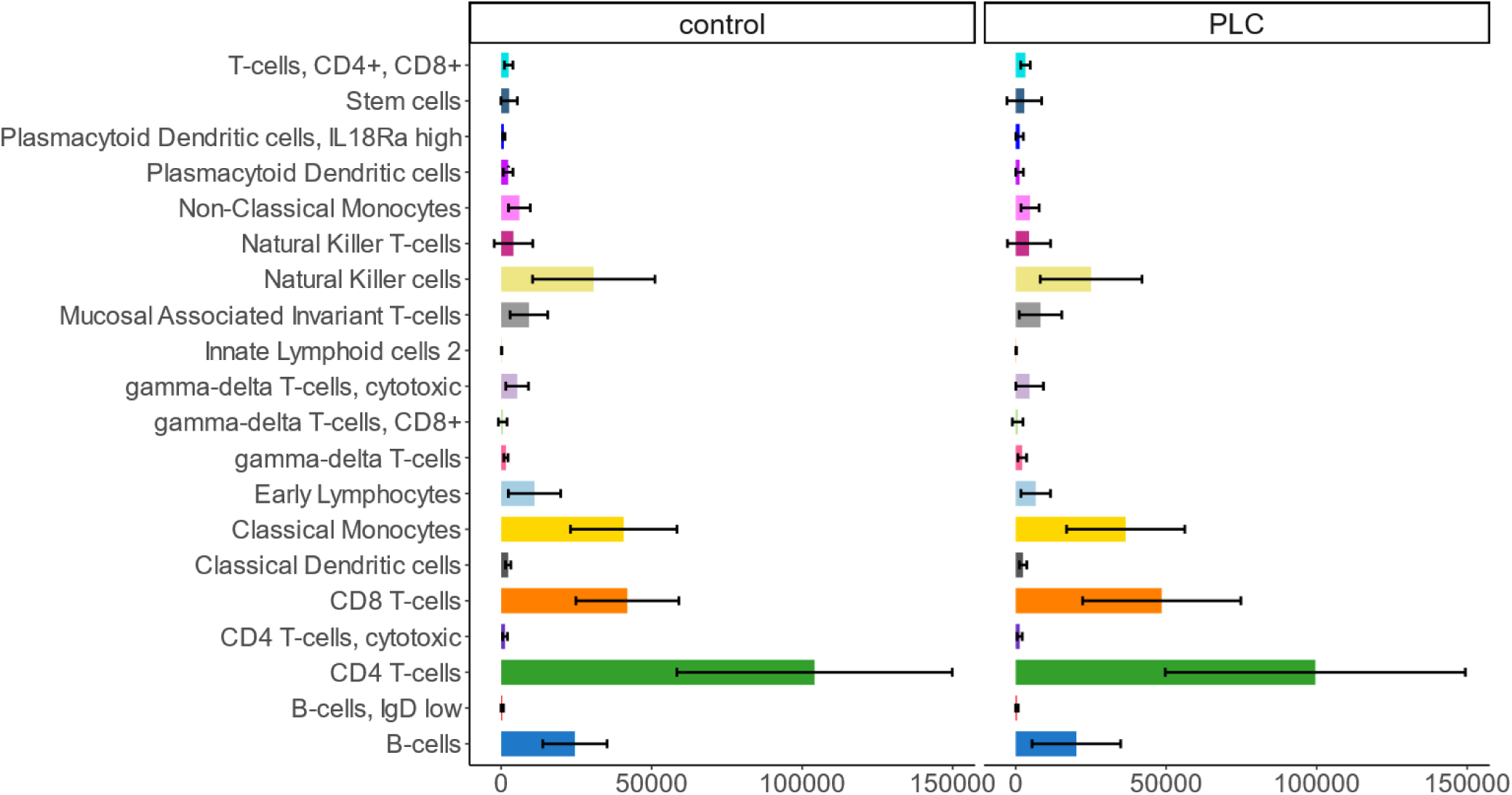
Composition plot showing the average absolute number of peripheral blood mononuclear cells and standard deviation characterized by CyTOF in control (left) and pulmonary long COVID (PLC, right) participants. Plasmacytoid dendritic cells were significantly reduced (p=0.048) in the PLC group.

**Supplemental Figure 8.**
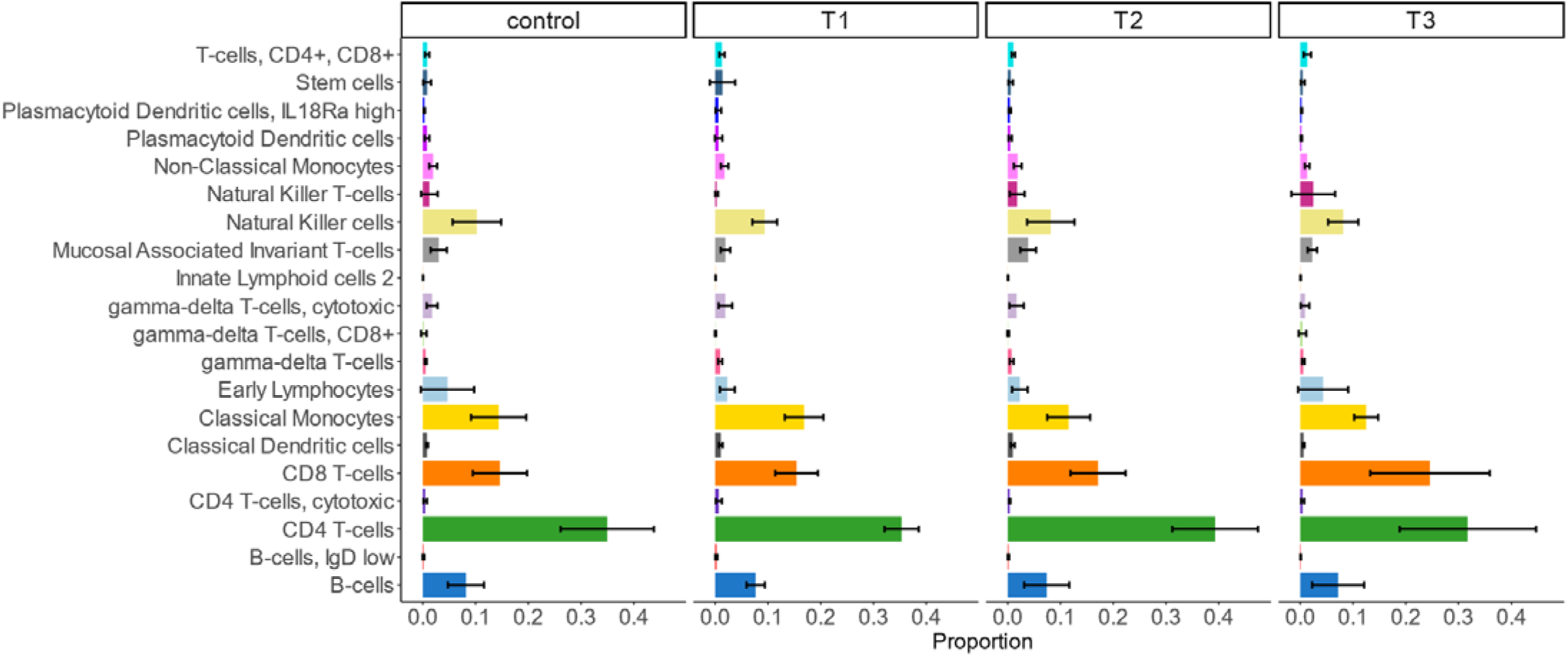
Composition plot of the average cell proportions and standard deviation of peripheral blood mononuclear cells from CyTOF by SGRQ score. PLC participants were divided by tertile, where T3 had the highest SGRQ scores, and T1 the lowest.

**Supplemental Figure 9.**
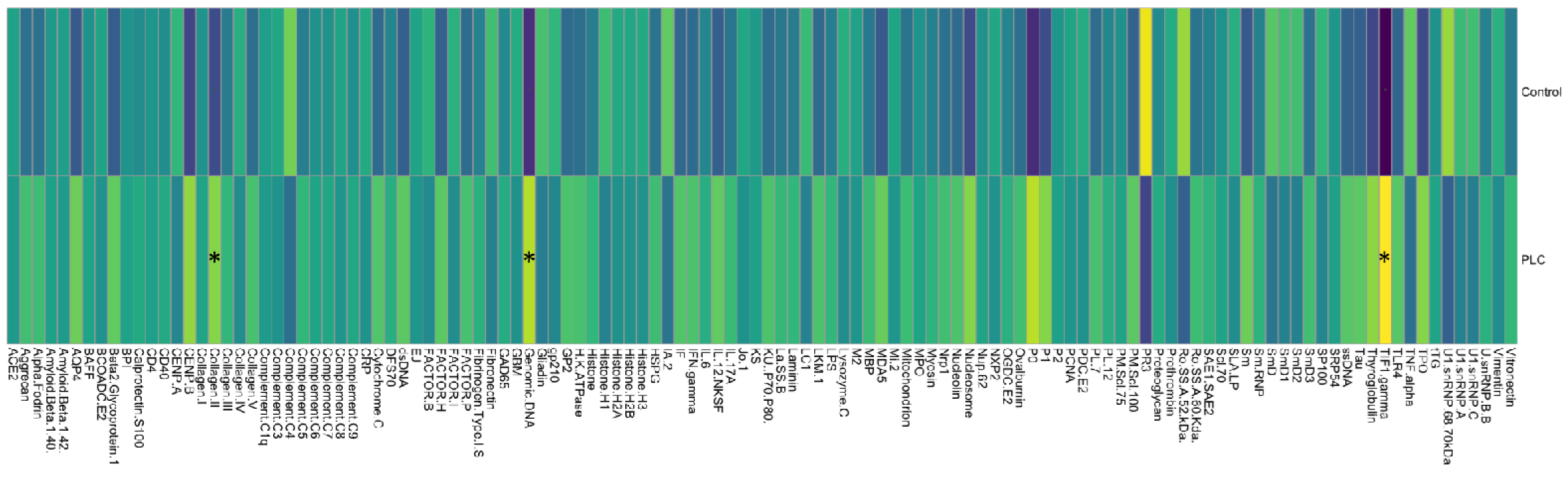
Heatmap of scaled normalized intensity signal of 120 auto IgG antibodies between PLC (bottom) and control (top) participants. Columns with * indicate p-value <0.05. Brighter colours indicate stronger presence of these antibodies. Notably, IgG antibodies against collagen II, genomic DNA and TIF-1 gamma were considered significant.

