## Supplementary Materials for "Single-cell multi-omics show disruption of blood and airway T-cells in pulmonary long COVID"

### *Tables*

#### **Table 1. Peripheral blood mononuclear cells antibody panel**

| Antigen | Conjugated metal* | Clone | Vendor | Cat. # |
| --- | --- | --- | --- | --- |
| CD38 | 106Cd | HIT2 | BioLegend | BL348202 |
| CD45RA | 110Cd | HI100 | BioLegend | BL304102 |
| CD45RO | 112Cd | UCHL1 | BioLegend | BL304202 |
| HLA-DR | 115Ln | L242 | BioLegend | BL307602 |
| IgD | 116Cd | IA6-2 | BioLegend | eBS12-5993-85 |
| CD8b | 141Pr | SIDI8BEE | eBioscience | TF14-5273-82 |
| CD19 | 142Nd | HIB19 | BioLegend | BL302202 |
| CD3e | 143Nd | OKT3 | BioLegend | BL317302 |
| a-KLRG1 | 144Nd | SA231A2 | BioLegend | BL367702 |
| CD31 | 145Nd | WM59 | BioLegend | BL303102 |
| CD11c | 147Sm | Bu15 | BioLegend | BL337202 |
| CD56 | 148Nd | NCAM16.2 | BD Bioscience | BD559043 |
| CD138 | 149Sm | DL-101 | BioLegend | BL352302 |
| CD116 | 150Nd | 4H1 | BioLegend | eTH14-1169-82 |
| CD14 | 151Eu | M5E2 | BioLegend | BL301802 |
| TCRgd | 152Sm | B1 | BioLegend | BL331202 |
| CD1c | 153Eu | L161 | BioLegend | BL331502 |
| CD301 (CLEC10A) | 154Sm | H037G3 | BioLegend | BL354702 |
| TCRab | 155Gd | T10B9.1A-31 | BD Bioscience | BD555546 |
| CD34 | 156Gd | 581 | BioLegend | BL343502 |
| CD16 | 158Gd | 3G8 | BioLegend | BL302002 |
| CD161 | 159Tb | HP-3G10 | BioLegend | BL339902 |
| CD32 | 160Gd | IV.3 | STEMCELL Technologies | SC60012 |
| CD94 (NKG2C) | 161Dy | DX22 | BioLegend | BL30550 |
| CD218a (IL-18Ra) | 162Dy | H44 | BioLegend | BL313804 |
| CD294 (CRTH2) | 163Dy | BM16 | BioLegend | BL350102 |
| CD123 | 164Dy | 6H6 | BioLegend | BL306002 |
| CD127 | 165Ho | A019D5 | BioLegend | BL351302 |
| CD8a | 168Er | SK1 | BioLegend | BL344702 |
| CD25 | 169Tm | BC96 | BioLegend | BL302602 |
| CD117 | 170Er | 104D2 | BioLegend | BL313202 |
| CD197 (CCR7) | 171Yb | G043H7 | BioLegend | BL353202 |
| CD304 (NRP1) | 172Yb | 12C2 | BioLegend | BL354502 |
| CD200R | 173Yb | 0X-108 | BioLegend | BL329302 |
| CD4 | 174Yb | SK3 | BioLegend | BL344602 |
| CD27 | 175Lu | O323 | BioLegend | BL302802 |
| FceRa1 | 176Yb | AER-37 | BioLegend | BL334602 |
| CD45 | 89Y | HI30 | BioLegend | BL304002 |

*Conjugation to metal was done by the UBC antibody lab (Vancouver, Canada)

#### **Table 2. Bronchoalveolar lavage antibody panel**

| Antigen | Conjugated metal* | Clone | Vendor | Cat. # |
| --- | --- | --- | --- | --- |
| CD1c | 153Eu | L161 | BioLegend | BL331502 |
| CD3e | 143Pr | OKT3 | BioLegend | BL317302 |
| CD4 | 174Yb | SK3 | BioLegend | BL344602 |
| CD8a | 168Er | SK1 | BioLegend | BL344702 |
| CD8b | 141Pr | SIDI8BEE | eBioscience | TF14-5273-82 |
| CD11c | 147Sm | Bu15 | BioLegend | BL337202 |
| CD14 | 151Eu | M5E2 | BioLegend | BL301802 |
| CD16 | 158Gd | 3G8 | BioLegend | BL302002 |
| CD19 | 142Nd | HIB19 | BioLegend | BL302202 |
| CD27 | 175Lu | O323 | BioLegend | BL302802 |
| CD38 | 106Cd | HIT2 | BioLegend | BL348202 |
| CD45 | 89Y | HI30 | BioLegend | BL304002 |
| CD45RA | 110Cd | HI100 | BioLegend | BL304102 |
| CD45RO | 112Cd | UCHL1 | BioLegend | BL304202 |
| CD56 | 148Nd | NCAM16.2 | BD Bioscience | BD559043 |
| CD94 (NKG2C) | 161Dy | DX22 | BioLegend | BL30550 |
| CD117 | 170Er | 104D2 | BioLegend | BL313202 |
| CD123 | 164Dy | 6H6 | BioLegend | BL306002 |
| CD127 | 165Ho | A019D5 | BioLegend | BL351302 |
| CD161 | 159Tb | HP-3G10 | BioLegend | BL339902 |
| CD197 (CCR7) | 171Yb | G043H7 | BioLegend | BL353202 |
| CD304 (NRP1) | 172Yb | 12C2 | BioLegend | BL354502 |
| FceRa1 | 176Yb | AER-37 | BioLegend | BL334602 |
| HLA-DR | 115Ln | L242 | BioLegend | BL307602 |
| IgD | 116Cd | IA6-2 | BioLegend | eBS12-5993-85 |
| TCRab | 155Gd | T10B9.1A-31 | BD Bioscience | BD555546 |
| TCRgd | 152Sm | B1 | BioLegend | BL331202 |
| CD163 | 173Yb | GHI/61 | Biolegend | 333602 |
| CD40 | 145Nd | 1C10 | Biolegend | 102802 |
| CD194 | 149Sm | MOPC-21 | Biolegend | 359402 |
| CD183 | 150Nd | G025H7 | Biolegend | 353733 |
| CD196 | 163Dy | G034E3 | Biolegend | 353427 |
| CD170 | 144Nd | 1A5 | Biolegend | 352002 |
| CD68 | 169Tm | KP1 | Biolegend | 916104 |
| CD80 | 162Dy | 2D10 | Biolegend | 305202 |
| CD86 | 160Gd | IT2.2 | Biolegend | 305435 |
| CD47 | 154Sm | CC2C6 | Biolegend | 323102 |

*Conjugation to metal was done by the UBC antibody lab (Vancouver, Canada)

#### **Table 3. Annotation guide for single-cell RNA sequencing and cytometry by time-of-flight**

| **Cell Type** | **Markers (single-cell RNA sequencing)** | **Markers (CyTOF)** |
| --- | --- | --- |
| CD4+ αβ T-cells | CD3e+, TCRAC+, TCRBC2+, IL7R+, CD40LG+ | CD3e+, CD4+, TCR-αβ+ |
| Cytotoxic CD4+ αβ T-cells | N/A | CD3e+, CD4+, TCR-αβ+, |
| CD8+ αβ T-cells | CD3e+, TCRAC+, TCRBC2+, CD8A+, CD8B+, CD2+, GZMB+ | CD3e+, CD8A+, CD8B+, TCR- αβ+ |
| Mucosal Associated Invariant T-cells | N/A | CD3e+, CD8A+, CD8B+, TCR- αβ+, IL18Rα+ |
| T-regulatory cells | CD3e+, TCRAC+, TCRBC2+, CD4+, FOXP3+, IKZF2+, CCR4+, CTLA4+ | CD3e+, CD4+, TCR-αβ+, CD25+ |
| Proliferating T-cells | CD3e+, CENPW+, TK1+, MKI67+ | N/A |
| Natural Killer T-cells | NKG7+, GNLY+, CD3e+ | CD3e+, TCR- αβ+, KLRG1+, CD56+ |
| CD4+, CD8+ αβ T-cells | N/A | CD3e+, CD8A+, CD8B+, CD4+, TCR- αβ+ |
| γδ T-cells | CD3e+, TRGC2+, TRDC+ | CD3e+, TCR-γδ+ |
| CD8+ γδ T-cells | CD3e+, TRGC2+, TRDC+, CD8A+, CD8+ | CD3e+, TCR-γδ+, CD8A+, CD8B+ |
| Cytotoxic γδ T-cells | N/A | CD3e+, TCR-γδ+, KLRG1+, CD94+, CD161+ |
| Natural Killer cells | NKG7+, GNLY+, CD3e- | KLRG1+, CD94+, CD56+, CD3e- |
| Innate Lymphoid cells 2 | N/A | CD117+, CD25+, CD127+, KLRG1+ |
| B-cells | MS4A1+ | CD19+, IgD+, HLA-DR+ |
| Early Lymphocytes | N/A | CD34 high |
| Stem cells | N/A | CD34 high, CD45 low |
| Dendritic cells | N/A | FceR1a high, HLA-DR+, CD11c+, |
| Dendritic cells 1 | XRC1+< CLEC9A+, CLNK+ | N/A |
| Dendritic cells 2 | FCER1G+, C1orf162+, CLEC7A+, PKIB+, CLEC10A+, CD1E+ | N/A |
| Migratory Dendritic cells | LAD1+, CCL19+ | N/A |
| Classical Dendritic cells | N/A | FceR1a high, CD1c+, HLA-DR+, CD11c+, CD123+ |
| Plasmacytoid dendritic cells | SCT+, SMPD3+, LILRA4+< BST2+, CLEC4C+, MAP3K2+, | FceR1a high, HLA-DR+, CD123+, CD1c-, CD11c- |
| Classical Monocytes | S100A12+ FCN1+, RNASE2+ | CD14+, HLA-DR+, CD32+, CD11c+ |
| Non-classical monocytes | MTSS1+, LILRA5+, FCN1+ | CD16+, HLA-DR+, CD32+ |
| Monocyte-derived macrophages | SPP1+, HAMP+, CCL2+ | HLA-DR+, CD1c+, CD11c+, CD86+ |
| Interstitial Macrophages perivascular | F13A1+, FOLR2+ | N/A |
| Alveolar Macrophages proliferating | TYMS+, CENPW+, MND1+ | N/A |
| Alveolar Macrophages metallothionein+ | CCL18+, MT1M+, MT1E+ | N/A |
| Macrophages CD68 low | N/A | CD86 high, CD80 low, CD68 low |
| Alveolar Macrophages M2 | CD163+, MRC1+ | CD163 high, CD40 low, CD86 high, CD80+ |
| Alveolar Macrophages M1 | CD80+, CD40 high, CD86+, FCGR1A+, FCGR3B+, FCGR2B+ | CD40 high, CD163 low, CD86 high, CD80+ |
| Alveolar Macrophages CCL3+ | CCL20+, FAM89A+ | N/A |
| Goblet cells | MUC5AC+, MUC5B+, KRT7+ | N/A |
| Ciliated cells | FOXJ1+, PIFO+, TPP3+, SNTN+ | N/A |

Abbreviations: Cytometry by time-of-flight (CyTOF)

#### **Table 4. Demographics of participants with CyTOF data of bronchoalveolar lavage**

All values are expressed as mean ± standard deviation unless otherwise specified.

|  | Pulmonary long COVID (n=4) | Control (n=3) | P-value |
| --- | --- | --- | --- |
| Age, years | 39.5 ± 5.9 | 34.7 ± 11.5 | 0.9 |
| Male, n (%) | 1 (25%) | 1 (33%) | 1 |
| BMI, kg/m^2^ | 24.1 ± 3.3 | 27.7 ± 2.3 | 0.1 |
| Tobacco smokers, n (%) |  |  | 0.4 |
| Former | 0 (0%) | 1 (33%) |  |
| Never | 4 (100%) | 2 (66%) |  |
| SGRQ, total | 51.1 ± 21.7 | 7.5 ± 5.5 | 0.06 |
| Symptoms | 52.1 ± 24.4 | 14.0 ± 6.1 | 0.06 |
| Activity | 52.1 ± 24.4 | 15.8 ± 13.7 | 0.2 |
| Impact | 67.8 ± 32.9 | 0.54 ± 0.94 | 0.05 |
| Pulmonary Function Test |  |  |  |
| FVC, L | 4.3 ± 1.0 | 4.7 ± 0.5 | 0.6 |
| FVC, % of predicted | 115.2 ± 13.0 | 118.8 ± 10.7 | 0.9 |
| FEV_1_, L | 3.5 ± 0.80 | 3.6 ± 0.42 | 1 |
| FEV_1_, % of predicted | 111.0 ± 6.8 | 110.1 ± 13.8 | 0.9 |
| FEV_1_/FVC | 80.4 ± 5.5 | 77.7 ± 4.3 | 0.6 |
| DLco, % of predicted | 118.8 ± 16.6 | 119.4 ± 6.6 | 0.4 |
| Blood cell counts |  |  |  |
| White blood cells (x 10^9^/L) | 5.4 ± 2.4 | 5.7 ± 0.5 | 0.6 |
| Neutrophils (%) | 52.5 ± 11.0 | 60.3 ± 1.6 | 0.2 |
| Lymphocytes (%) | 38.3 ± 9.9 | 29.6 ± 2.5 | 0.1 |
| Monocytes (%) | 5.0 ± 2.2 | 5.3 ± 1.5 | 0.9 |
| Eosinophils (%) | 1.3 ± 0.34 | 1.6 ± 0.5 | 0.4 |
| Bronchoalveolar Lavage |  |  |  |
| Total Nucleated cells | 138.3 ± 58.0 | 365.2 ± 202.6 | 0.2 |
| Alveolar Macrophage (%) | 85.1 ± 10.9 | 71.8 ± 27.6 | 0.6 |
| Lymphocytes (%) | 10.1 ± 7.1 | 15.5 ± 12.6 | 0.6 |
| Neutrophils (%) | 3.8 ± 3.9 | 11.0 ± 14.3 | 0.3 |
| Eosinophils (%) | 0.0 ± 0.0 | 1.5 ± 0.87 | 0.03 |
| Bronchial lining cells (%) | 1.0 ± 1.4 | 0.0 ± 0.0 | 0.3 |
| COVID-19 Severity, n (%) |  |  | 0.7 |
| Hospitalized | 0 | 0 (0%) |  |
| Mild | 4 (100%) | 2 (66%) |  |
| Unexposed | NA | 1 (33%) |  |
| Computed Tomography, n |  |  |  |
| Bronchiectasis | 2 (50%) | 0 (0%) | 0.26 |
| Consolidations | 0 (0%) | 0 (0%) | 1 |
| Ground Glass Opacities | 0 (0%) | 0 (0%) | 1 |
| Honeycombing | 0 (0%) | 0 (0%) | 1 |
| Linear scars or atelectasis | 2 (50%) | 0 (0%) | 0.26 |
| Mosaic attenuation | 0 (0%) | 0 (0%) | 1 |
| Reticulations | 1 (25%) | 0 (0%) | 0.56 |
| Emphysema | 0 (0%) | 0 (0%) | 1 |

Abbreviations: BMI (body mass index), SGRQ (Saint George’s Respiratory Questionnaire) FVC (forced vital capacity), FEV1 (forced expiratory volume in one second), DLco (diffusion capacity for carbon monoxide)

#### **Table 5. Demographics of participants with serum autoantibody data**

All values are expressed as mean ± standard deviation unless otherwise specified.

|  | Pulmonary long COVID (n=22) | Control (n=18) | P-value |
| --- | --- | --- | --- |
| Age, years | 44.0 ± 12.9 | 42.2 ± 19,1 | 0.61 |
| Male, n (%) | 10 (45%) | 9 (50%) | 0.78 |
| BMI, kg/m^2^ | 26.0 ± 4.3 | 26.5 ± 4.3 | 0.78 |
| Tobacco smokers (%) |  |  | 0.41 |
| Current | 1 (4.5%) | 0 (0%) |  |
| Former | 5 (23%) | 3 (17%) |  |
| Never | 16 (72.5) | 15 (83%) |  |
| SGRQ, total | 47.6 ± 17.4 | 4.2 ± 3.6 | 7.9 10^-8^ |
| Symptoms | 55.1 ± 17.6 | 8.1 ± 15.0 | 1.0 10^-7^ |
| Activit y | 63.1 ± 23.5 | 7.5 ± 8.7 | 1.2 10^-7^ |
| Impact | 36.0 ± 19.1 | 1.0 ± 1.8 | 5.5 10^-8^ |
| Pulmonary Function Test |  |  |  |
| FVC, L | 4.3 ± 0.95 | 4.6 ± 0.85 | 0.51 |
| FVC, % of predicted | 108.0 ± 18.5 | 116 ± 10 | 0.13 |
| FEV_1_, L | 3.4 ± 0.72 | 3.6 ± 0.66 | 0.25 |
| FEV_1_, % of predicted | 103.8 ± 18.3 | 113 ± 12 | 0.16 |
| FEV_1_/FVC | 78.9 ± 7.9 | 79.6 ± 7.1 | 0.93 |
| DLco, % of predicted | 102.8 ± 17.0 | 114 ± 15 | 0.06 |
| Blood cell counts |  |  |  |
| White blood cells (x 10^9^/L) | 6.2 ± 1.8 | 5.3 ± 1.1 | 0.18 |
| Neutrophils (%) | 59.4 ± 9.0 | 58.7 ± 7.9 | 0.45 |
| Lymphocytes (%) | 31.0 ± 8.6 | 30.6 ± 6.8 | 0.94 |
| Monocytes (%) | 5.5 ± 1.5 | 4.9 ± 0.90 | 0.23 |
| Eosinophils (%) | 1.8 ± 0.84 | 2.5 ± 1.6 | 0.11 |

Abbreviations: BMI (body mass index), SGRQ (Saint George’s Respiratory Questionnaire) FVC (forced vital capacity), FEV1 (forced expiratory volume in one second), DLco (diffusion capacity for carbon monoxide)
